# Microcirculation Mirror: A zero-install browser workstation for hand microvascular anatomy and nailfold capillaroscopy

**DOI:** 10.64898/2026.11.15.26368085

**Authors:** Yingyu Wang, Jingjing He, Weiqi Liu, Xiang Xu, Bingwei Li, Yuan Li, Bing Wang, Mingming Liu

## Abstract

**Background:** Hand microvasculature is conventionally presented through diagrams, cadaveric atlases and specialized optical instruments, while conventional clinical radiography cannot resolve individual capillary loops (5-15 µm lumen) in vivo, which dominate cutaneous oxygen exchange.

**Methods:** We present *Microcirculation Mirror*, a zero-install browser workstation that registers layered hand anatomy to a live 21-landmark pose and couples it to a nailfold video capillaroscopy (NVC) simulator, a lumped microcirculatory model and an imaging-physics module. The single-file application runs offline on a laptop or, over local HTTPS, on a phone as an installable app.

**Results:** Its 3D atlas contains 136 named structures per hand in six systems from open Z- Anatomy/BodyParts3D meshes, plus a literature-parameterized capillary layer. The NVC module renders a 200×, 1.2-mm field with six deterministic patterns and quantified density, giant capillaries, avascular width fraction, hemorrhages, tortuosity and ramification. A lumped model reports nine estimates (SIM); a physics module shows that a 10-µm capillary occupies 0.07 of a 150-µm detector pixel; with iodinated contrast it retains ≈0.016% effective contrast, about 60× below a 1% detection threshold; unenhanced blood is ≈270× lower. The 2D stage renders in 2.9 ms per frame; system merging reduces atlas draw calls from 136 to 6.

**Conclusions:** The camera-overlay layers are templates, the atlas macro-anatomy is open mesh data, and the capillary layer is sampled. No patient data are used. The instrument is a product prototype, not a medical device, and provides a reproducible framework for making multi-scale microvascular reasoning visible.

## Introduction

The microcirculation of the hand is studied across rheumatology, physiology and anatomy. Rheumatologists examine nailfold capillary loops to identify the scleroderma pattern and distinguish its early, active and late stages [1–3]. Physiologists investigate oxygen transport from capillaries measuring 5 to 15 µm in diameter to the surrounding tissue [4–6], while anatomists characterize the palmar arches and digital arteries supplying the skin [7, 8]. Despite these complementary perspectives, key microvascular structures and features, including capillary loops, the papillary plexus and the distinction between normal and abnormal nailfold architecture, remain difficult to visualize. Individual capillaries cannot be resolved using conventional clinical radiography [9, 10], are generally inaccessible to surface microscopy outside the nailfold, and exhibit variability that cannot be adequately represented by a single static diagram.

These limitations reflect a broader challenge in integrating anatomical information across spatial scales and imaging modalities. Cadaveric dissection reveals macroscopic vascular anatomy but does not readily demonstrate individual capillary loops. Two-dimensional atlases typically present the palmar arterial system and microvascular images separately, with limited spatial correspondence between them. Clinical nailfold video capillaroscopy provides magnified views of the living nailfold, but its use depends on dedicated equipment, operator expertise and standardized acquisition procedures, including the four- field protocol considered here. Consequently, conventional anatomical resources and clinical capillaroscopy offer limited support for continuous exploration from a digital artery to the capillary bed it supplies.

Computational anatomy provides a potential framework for connecting these scales. Open mesh collections, including BodyParts3D and Z-Anatomy, contain named three-dimensional models of the hand and forearm [11,12]. Browser-based hand tracking can estimate a pose comprising 21 landmarks from a single RGB frame [13], while Canvas2D and WebGL support the real-time rendering of layered anatomical annotations. Integrating these capabilities into a coherent interactive interface also requires clear differentiation among measured structures, anatomical templates, sampled features and simulated phenomena. Recent preprints describing browser-based anatomy instruments provide related examples at comparable levels of evidence in anatomy and biomedical visualization [14, 15].

Microcirculation Mirror was developed to investigate whether a browser-based instrument could combine these capabilities within a single HTML file while preserving anatomical relationships and explicitly communicating the limitations of its visual representations. The application registers skin, muscle, tendon, bone, nerve, artery, capillary and vein layers to a tracked hand and provides a separate three-dimensional hand atlas derived from open meshes. It also renders a simulated nailfold video capillaroscopy field at 200-fold magnification with six selectable patterns, implements a lumped microcirculatory model, and uses attenuation and detector calculations to examine whether conventional clinical X-rays can resolve individual hand capillaries in vivo. The application is designed to operate offline on a laptop or mobile phone without installation or reliance on a server-side database.

Our work presents the architecture and technical characterization of *Microcirculation Mirror* as a research and demonstration prototype. Its contributions comprise a reproducible approach to pose- registered, multiscale microvascular anatomy in the browser, an open quantitative hand atlas that distinguishes mesh-derived anatomy from procedural microanatomy, and integrated simulation and imaging-physics modules that clarify both the basis and the limitations of the visualized information. Evaluation is based on technical characterization and worked examples, consistent with the scope of a methods and instrument description.

## Results

### A single-file instrument for multi-scale hand anatomy

The application is distributed as a single HTML file with embedded CSS and JavaScript and support for optional local assets, including a float16 hand-landmark model, six GLB layers per hand, and a compact Three.js build. It requires no runtime build step, package manager, or server-side component. The same file supports four operating modes, comprising a demonstration with an illustrative hand, camera-based tracking in which MediaPipe HandLandmarker estimates 21 landmarks [13], a full-screen three-dimensional atlas, and a microscopy and imaging-physics workspace (Fig. 1).

**Fig. 1.**
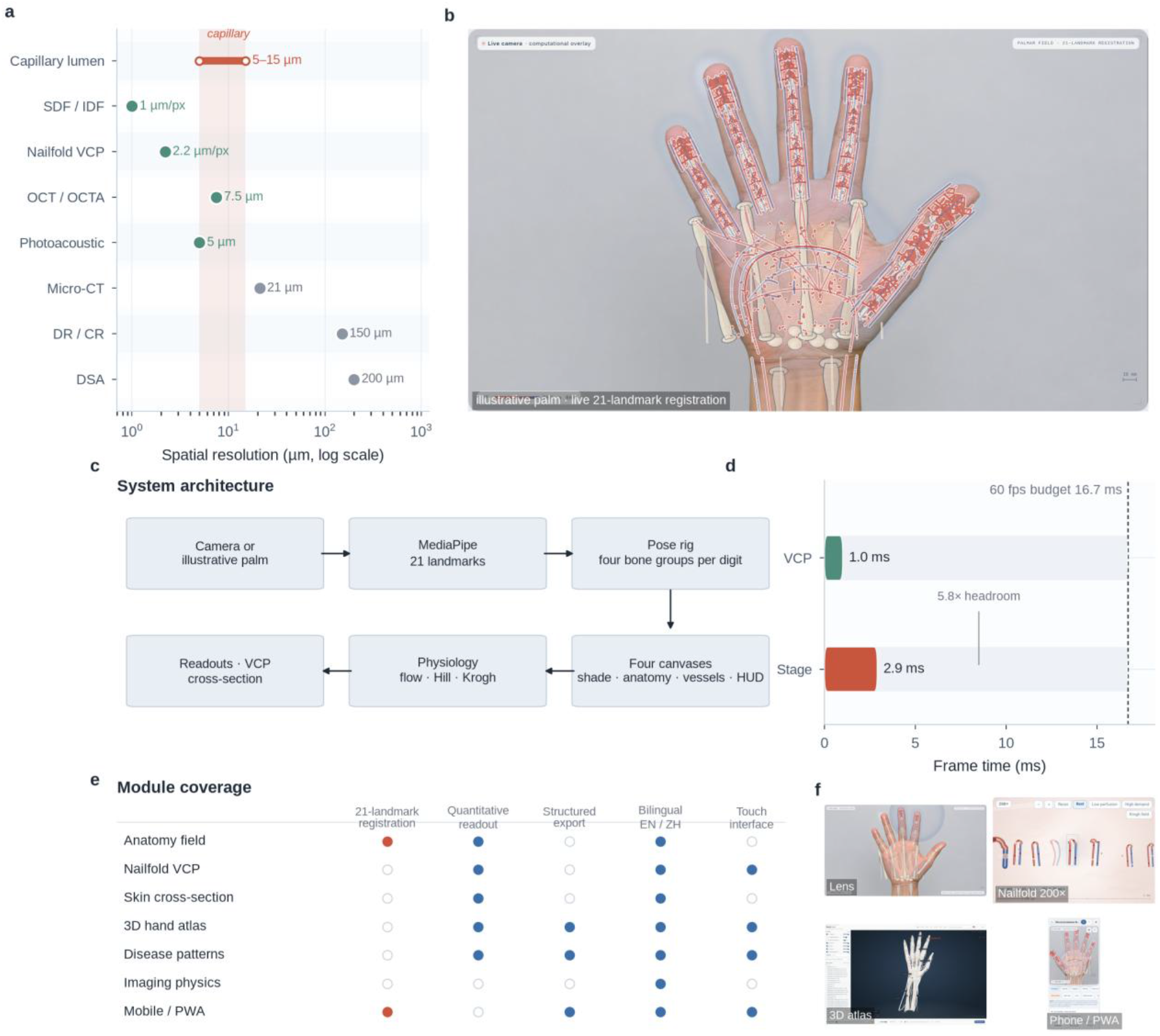
A zero-install instrument for multi-scale hand microvascular anatomy. **a**, Spatial resolution of imaging modalities on a logarithmic scale. The shaded band marks the 5–15-µm capillary lumen; optical modalities (SDF/IDF, NVC, OCT, photoacoustic) reach it, while conventional radiography, DSA and laboratory micro-CT do not in routine clinical use. **b**, The application running on an illustrative palm: a 21-landmark pose registers skin, bone, muscle, tendon, nerve, artery, capillary and vein layers in real time. **c**, System architecture. Camera input or an illustrative palm provides the basis for a 21-landmark pose represented in the Media Pipe format. A pose rig comprising the palm, four bone groups per digit, and the forearm supports rendering across four canvases for shading, anatomy, vessels, and the heads-up display (HUD). A physiology model incorporating flow, Hill, and Krogh formulations supplies model- derived readouts alongside the nailfold video capillaroscopy (NVC) and cross-sectional views. **d**, Frame budget. The 2D stage renders in 2.9 ms and the nailfold field in 1.0 ms against the 16.7-ms budget for 60 fps. **e**, Module coverage matrix. **f**, Thumbnails of the lens reveal, nailfold field, 3D atlas and phone layout. All screenshots are from the application. SIM indicates model estimates.

The interface maintains the hand as a shared spatial reference while allowing users to change anatomical layers and viewing scales. The main stage presents the hand, while a panel on the right provides controls for anatomical layers, visualization settings, physiology, and numerical readouts. Presets provide access to arterial, capillary, venous, musculoskeletal, and nailfold views. Additional viewing controls allow users to alternate among a whole-field view, a split view, a lens centered on the index fingertip, a depth section, and a nailfold lens. Selecting a visible vessel opens an inspector containing its anatomical name, vessel class, lumen diameter, depth, erythrocyte velocity, oxygen saturation (SO₂), wall shear stress, and segment length.

Rendering is organized into four coordinated canvases (Fig. 1c). The shading canvas renders the skin envelope or camera veil, and the anatomy canvas renders bones, muscles, tendons, nerves, and lymphatics. A WebGL or Canvas2D canvas renders arteries, capillaries, veins, and erythrocytes, while an annotation canvas presents labels, the viewing lens, scale bars, and the depth ruler. All four canvases use the same landmark-derived pose, allowing the rendered layers to move together as hand position changes. A lumped physiology model uses the shared application state to generate the nine numerical readouts presented in the control panel.

## Pose-registered macro-anatomy

The tracker estimates 21 landmarks in screen coordinates, from which the application constructs orthogonal coordinate frames for the palm, four bone groups per digit, and the forearm. The digital groups comprise 20 groups in total, with the thumb represented using its carpometacarpal-to- metacarpophalangeal segment. These frames are scaled according to tracked joint distances and smoothed using a frame-rate-aware exponential filter. Temporary tracking losses are bridged for up to 18 frames before the overlay is cleared. The resulting anatomical representation is pose-registered rather than segmented from the camera image. Bones, muscles, tendons, and nerves are canonical templates whose geometry is derived from hand proportions (Fig. 2).

**Fig. 2.**
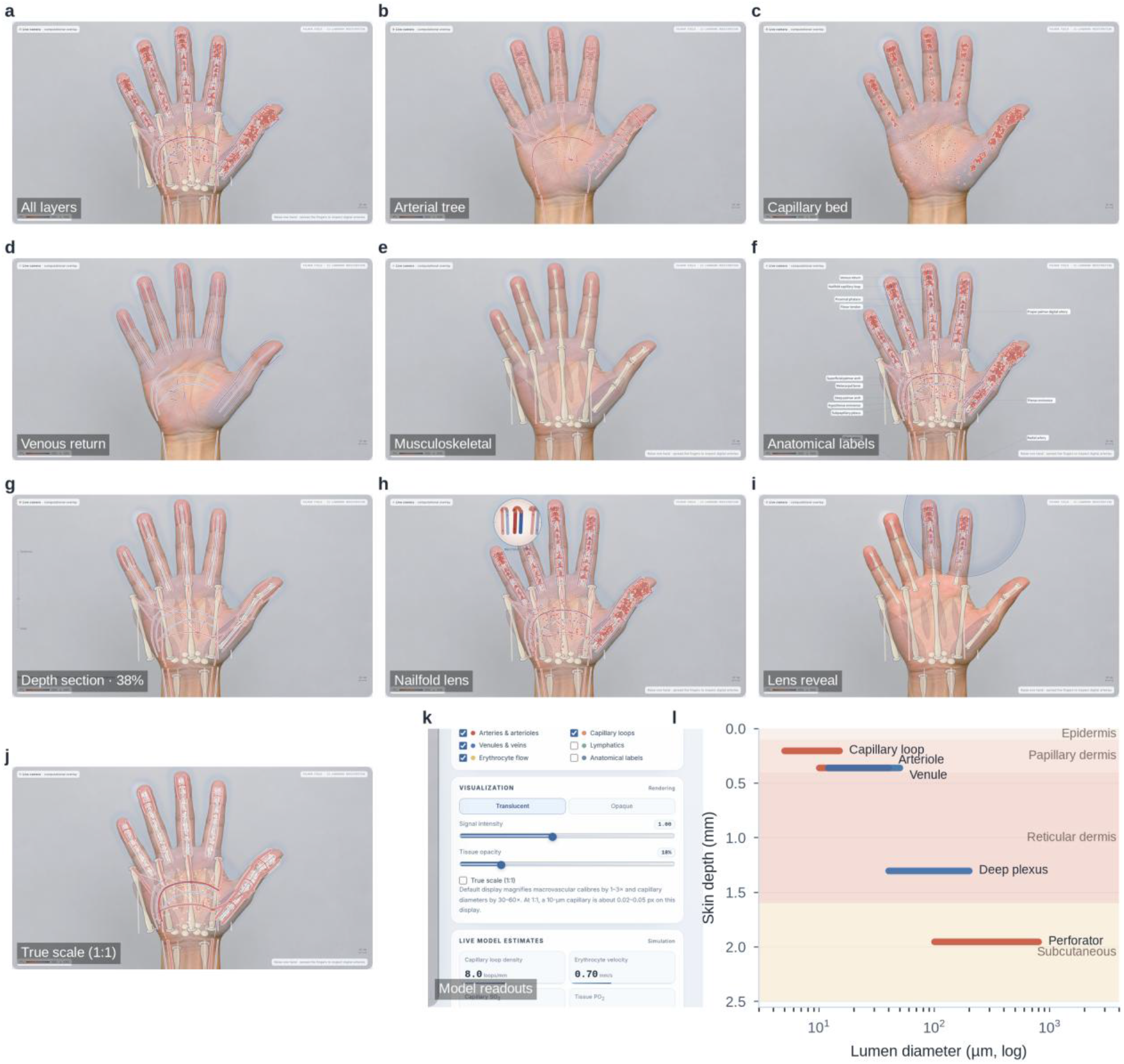
Pose-registered anatomical layers on an illustrative palm. **a**, All layers. **b**, Arterial tree. **c**, Capillary bed. **d**, Venous return. **e**, Musculoskeletal layer. **f**, Anatomical labels. **g**, Depth section at 38%. **h**, Nailfold lens. **i**, Lens reveal around the index fingertip. **j**, True scale (1:1), in which a 10-µm capillary is 0.02–0.05 display px. **k**, Model readout panel. **l**, Depth–calibre map of the skin microvasculature.

Anatomical layers can be viewed independently or in combination. The arterial layer extends from the radial and ulnar arteries through the superficial and deep palmar arches to the common and proper palmar digital arteries, arterioles, and capillary loops. The venous layer includes venules, dorsal digital veins, and the superficial and deep palmar venous networks. The musculoskeletal layer represents the thenar and hypothenar eminences, interossei, lumbricals, and flexor tendons, while the neural layer depicts schematic median and ulnar nerve trunks and their digital branches. An optional lymphatic layer provides a sparse, blind-ended network. The skin envelope can be rendered as either a translucent veil or an opaque silhouette. A depth-section view attenuates structures according to their modeled depth, while a circular lens centered on the index fingertip reveals the combined anatomical layers.

The vessel inspector provides quantitative information for individual vascular segments, including modeled lumen diameter, depth, erythrocyte velocity, oxygen saturation (SO₂), and wall shear stress. Wall shear stress is calculated from the Poiseuille velocity profile as *τ* = 8*ηv*/*D*, with viscosity set to *η* = 0.0035 Pa s. All values are model estimates, and the inspector allows users to examine differences across vascular scales, including the nearly two-order-of-magnitude difference in diameter between a 0.9-mm proper palmar digital artery and a 10-µm capillary, together with their corresponding modeled velocities.

The true-scale view relates anatomical dimensions to the magnification required to make small structures visible. In the default view, macrovascular diameters are shown at approximately 1–3 times their modeled size and capillary diameters at 30–60 times their modeled size, while the capillary bed is sampled well below its expected anatomical density. At a scale of 1:1, a 10-µm capillary occupies approximately 0.02–0.05 pixels on a typical screen and therefore becomes a subpixel structure. The resulting visualization illustrates a sampling limitation analogous to the detector-sampling constraints that, together with limited attenuation contrast, prevent conventional clinical radiography from resolving individual capillary loops in vivo.

## An open-mesh 3D atlas with a procedural capillary layer

The 3D atlas is a separate full-screen viewer built from the Z-Anatomy collection, with geometry derived from BodyParts3D [16]. The hand subset comprises 136 named structures per hand across six anatomical systems, including 29 bones, 14 joint capsules and ligaments, 47 muscles, fasciae, and tendon sheaths, 13 arterial segments, 10 venous segments, and 23 nerve segments (Fig. 3). The skeletal structures include the radius, ulna, eight carpals, five metacarpals, and fourteen phalanges. Each structure is annotated with English and Chinese names and mesh statistics (Supplementary Fig. 6); 36 structures also have a Foundational Model of Anatomy (FMA) identifier, and 26 include a brief description. The left and right hands are extracted independently.

**Fig. 3.**
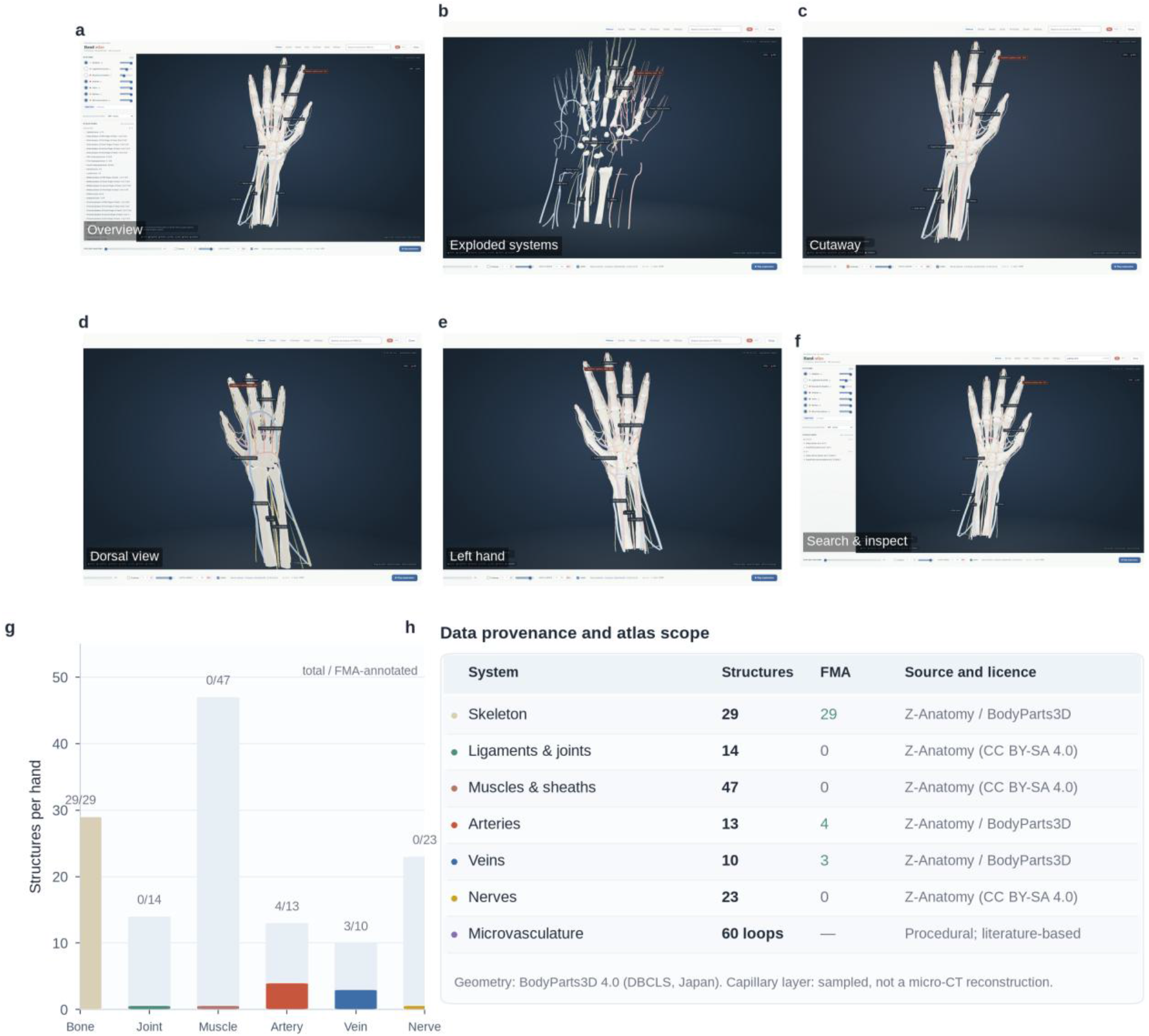
Open-mesh 3D hand atlas and its provenance. **a**, Atlas overview with the structure tree and layer controls. **b**, Exploded systems. **c**, Cutaway. **d**, Dorsal view. **e**, Left hand. **f**, Search and inspect. **g**, Structure inventory (total bars) and FMA-annotated structures (coloured bars). **h**, Provenance and scope of each system. The 136 structures per hand are derived from Z-Anatomy/BodyParts3D; the 60-loop capillary layer is procedural and sampled for legibility.

The atlas contains seven independently controlled layers representing the skeleton, ligaments and joints, muscles and sheaths, arteries, veins, nerves, and procedural microvasculature. Visibility and opacity can be adjusted for each layer. A two-stage exploded-view control first separates anatomical systems and then individual structures, while a cutaway plane clips the model along the X, Y, or Z axis. Search supports English names, Chinese names, and FMA identifiers. Selecting a structure opens an inspector showing its name, anatomical system, FMA identifier, bounding-box dimensions, and mesh counts. Seven viewing presets, palmar, dorsal, radial, ulnar, proximal, distal, and oblique, and a 26- second guided tour provide reproducible viewpoints. Switching between hands loads the corresponding set of six GLB files.

Neither source dataset includes capillary, arteriole, or venule meshes. The microvascular layer is therefore generated at runtime on the volar surfaces of the five distal phalanges, with approximately 12 loops per fingertip and 60 loops per hand. Loop geometry is informed by published morphometry, including a loop height of 278.5 ± 53.3 *μ*m and limb diameters of 5.6–13.4 µm from volar-digit measurements [17], together with an outer loop width of approximately 39 µm [18]. The layer supports magnifications of 1, 10, and 20 times and is labeled “sampled for legibility”. At unit magnification, loop height and width follow the published dimensions and become subpixel at the viewing scale used, whereas tube diameter remains exaggerated at all magnifications to preserve visibility. The resulting geometry is an illustrative model.

Provenance is documented throughout the atlas. Macro-anatomical structures use reference mesh data, whereas microvascular structures are procedurally generated. Their respective origins are identified in the layer labels, structure inspector, and methods note, with supporting documentation in the repository (Supplementary Fig. 1 and Supplementary Table S1). The atlas combines reference anatomy and synthesized microvascular geometry within a shared environment while retaining visible information about the source and status of each layer.

## Nailfold video capillaroscopy at 200×

The microscopy workspace renders a 200× nailfold videocapillaroscopy field with 1 mm = 500 px and a 1.2-mm field width (Fig. 4). Each loop is drawn as a hairpin tube with an arterial limb, a venous limb, an apical segment and a pale lumen; erythrocyte particles move at a nominal ×0.1 slow motion, decoupled from the reported velocity. A 1-mm ruler and a 200-µm sub-scale are always visible. The user can select a loop, read its number, width and erythrocyte velocity, and switch between hairpin, tortuous, crossed and branched morphologies. A Krogh field can be overlaid on the selected apex, with a radial PO₂ field and a PO_2_(*r*) curve. A separate skin cross-section shows the epidermis, papillary dermis, subpapillary plexus, reticular dermis, deep dermal plexus and subcutaneous tissue over 0–2.5 mm, with capillary loops, an arteriole, a venule, a lymphatic and the Krogh field.

**Fig. 4.**
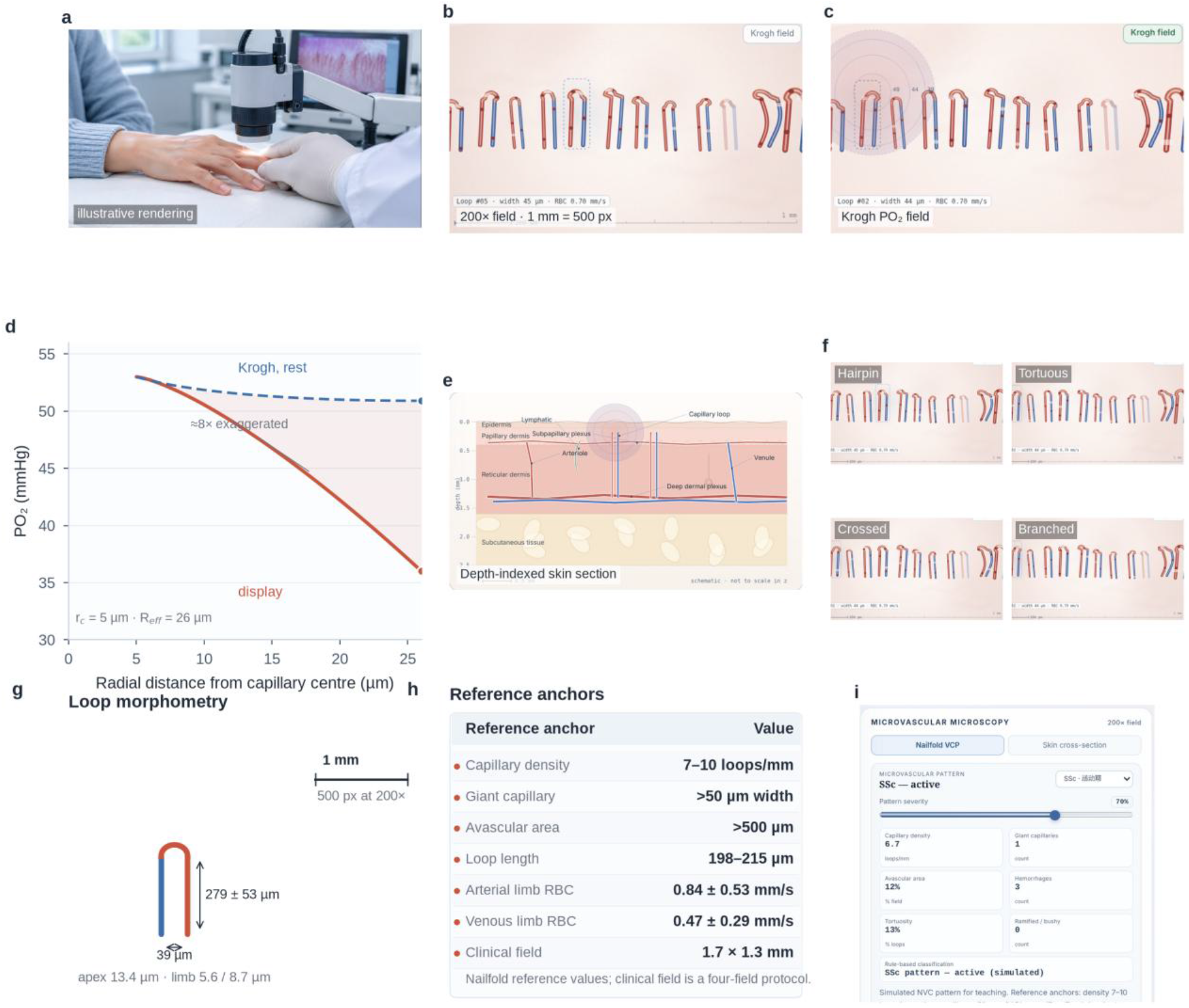
Nailfold video capillaroscopy module. **a**, Nailfold examination (illustrative rendering). **b**, 200× field with a 1-mm ruler (1 mm = 500 px). **c**, Krogh PO₂ field around the selected loop. **d**, Normalised display PO_2_(*r*) and the physical Krogh solution at rest (*M* = 5 × 10^−4^ cm^3^ O₂ cm^−3^ s^−1^). **e**, Depth- indexed skin cross-section (0–2.5 mm). **f**, Hairpin, tortuous, crossed and branched loop morphologies. **g**, Loop morphometry and calibration. **h**, Reference anchors. **i**, Rule-based pattern report in the interface. The field is a single illustrative 1.2-mm window; morphology and flow are simulated.

The module uses published reference values for normal capillary density of 7–10 loops/mm, loop length of 198–215 µm, outer loop width of approximately 39 µm, and limb diameters of 5.6–13.4 µm [17, 18]. Reference erythrocyte velocities are 0.84 ± 0.53 mm/sin the arterial limb and 0.47 ± 0.29 mm/sin the venous limb [19], and the giant-capillary threshold is set at 50 µm. The simulated field represents a single illustrative 1.2-mm window rather than the four-field clinical acquisition protocol. The overlaid Krogh field is a normalized approximation for visualization; the analytical Krogh solution is presented separately in the Methods and Fig. 4d for comparison.

## Six deterministic microvascular patterns

The pattern panel and the 3D capillary layer share six simulated profiles representing healthy microvasculature, nonspecific Raynaud-associated changes, early, active, and late systemic sclerosis (SSc) patterns, and diabetes-associated changes (Fig. 5). Each profile defines capillary density, the fractions of giant capillaries and avascular regions, hemorrhage count, tortuosity, and the fraction of ramified or bushy loops. A severity slider interpolates between the healthy baseline and the selected profile, and selecting a disease-associated pattern sets the slider to its nominal severity. Pattern generation is deterministic, with the selected profile, severity, and fixed seed producing the same field on repeated rendering. Users can therefore reproduce individual fields and compare profiles without variation from random resampling.

**Fig. 5.**
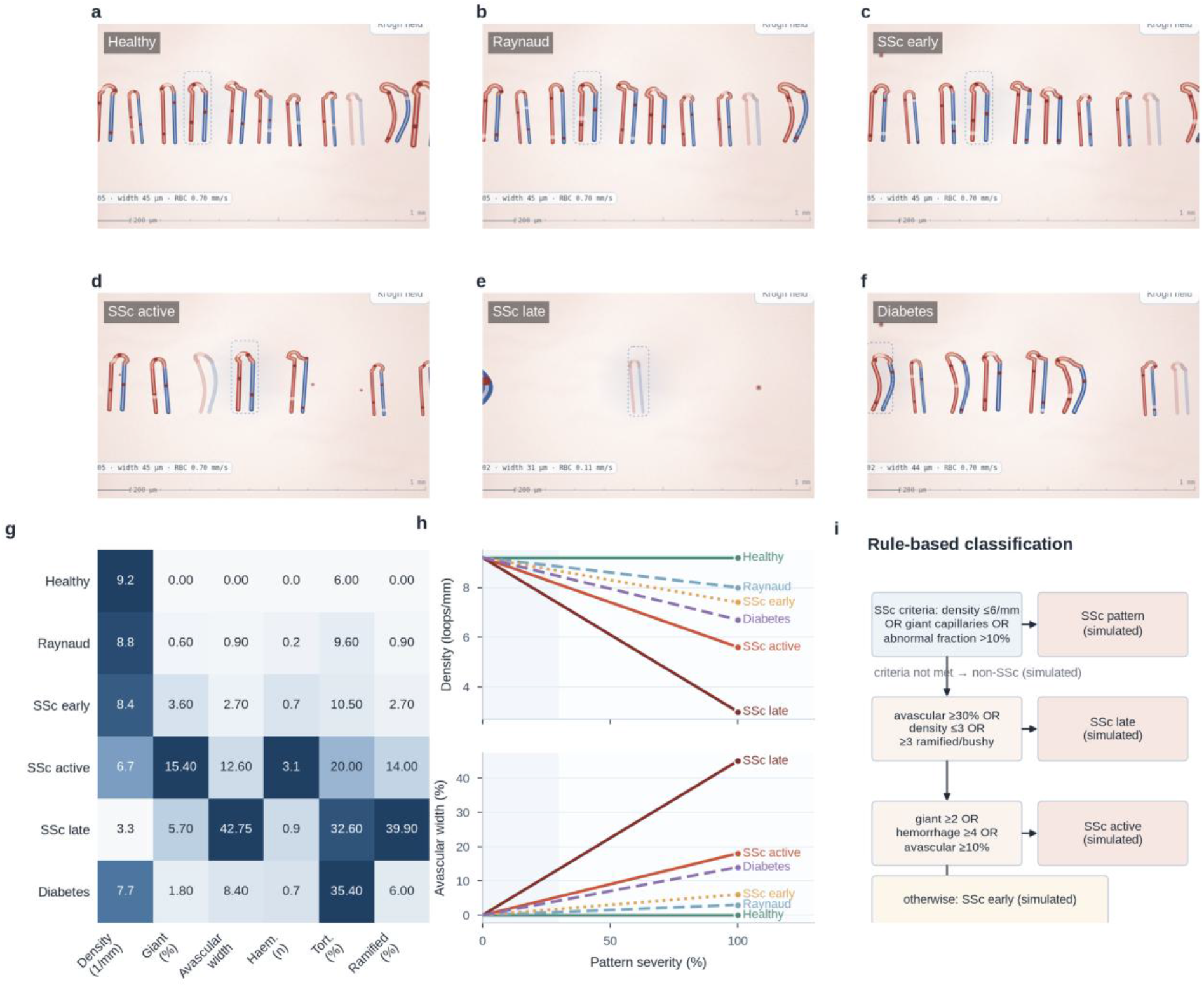
Six deterministic microvascular patterns. **a**–**f**, Healthy, Raynaud non-specific, SSc early, SSc active, SSc late and diabetes fields, each rendered at its nominal severity. **g**, Feature heatmap of the six profiles at nominal severity. **h**, Severity-blend curves for density and avascular width fraction. **i**, Rule- based classification flow inspired by Fast Track and CAPI-score.

The healthy profile uses a visualization density of 9.2 loops/mm to generate the simulated field, whereas the physiology panel uses a separate default lumped estimate of 8.0 loops/mm. The pattern panel reports seven metrics, including capillary density (loops/mm), giant-capillary count, avascular width fraction (% of field width), expected hemorrhage count, tortuosity (% of loops), ramified or bushy loop count, and the fraction of abnormal loops. A rule-based classifier inspired by the Fast Track algorithm and the CAPI-score assigns labels of “normal”, “non-specific”, “SSc pattern—early”, “SSc pattern— active”, “SSc pattern—late”, or “diabetes-associated microangiopathy”, each accompanied by the qualifier “simulated” [20, 21]. The classification rules are simplified for illustration and do not constitute a validated implementation of either method. Automated nailfold videocapillaroscopy (NVC) analysis remains an active area of research [22].

The Raynaud and diabetes profiles include small giant-capillary fractions as visualization parameters. These parameters are not used to assign an SSc classification to either profile. The panel allows users to inspect and quantify the morphological features underlying the simulated classifications. The heatmap summarizes differences in density, giant-capillary count, avascular width fraction, hemorrhages, tortuosity, and ramification across the six profiles (Fig. 5g), while the severity curves show how these features vary between the healthy baseline and each disease-associated profile. A structured JSON report can be exported with the selected pattern, severity, metrics, reference ranges, and a disclaimer stating that the output is not intended for diagnosis (Supplementary Fig. 4).

## A lumped physiology model with uncertainty

The physiology panel provides six adjustable inputs, including mean arterial pressure (MAP), skin temperature, sympathetic tone, capillary recruitment, oxygen demand, and animation speed. These inputs are mapped to nine readouts through a lumped model (Table 1). Capillary recruitment adjusts the number of loops represented as perfused in the visualization and does not imply a change in anatomical capillary density. The flow index is

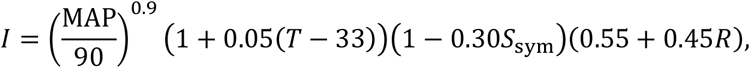

and blood flow is *Q* = clamp(6.4*I*, 0.8,12) mL/100 g/min. Erythrocyte velocity follows a Poiseuille-like scaling,

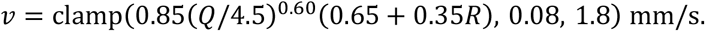

Oxygen saturation follows the Hill equation with *P*_50_ = 26.6 mmHg and *n* = 2.7 [5,23]; oxygen extraction is *O*_2_*ER* = clamp(0.23 *VO*_2_(4.5/*Q*), 0.08,0.72) ; and tissue PO₂ is approximated as 0.70*P_v_O*_2_ + 8 mmHg. The microvascular flow index and heterogeneity index are empirical adaptations from the sublingual microcirculation literature [24,25]; in the hand model they are approximations, not the consensus definitions, and both are clamped to their stated ranges.

**Table 1.** Default model estimates and units.

| **Readout** | **Default** | **Unit** |
|---|---|---|
| Capillary loop density | 8.0 | loops/mm |
| Erythrocyte velocity | 0.70 | mm/s |
| Capillary SO<sub>2</sub> | 86 | % |
| Tissue PO<sub>2</sub> | 36 | mmHg |
| Microvascular flow index (MFI) | 3.0 | 0–3 |
| Heterogeneity index (HI) | 21 | % |
| Oxygen extraction ratio | 23 | % |
| Blood flow | 4.5 | mL/100 g/min |
| Capillary transit time | 0.85 | s |

All values are SIM model estimates.

At the defaults (MAP 90 mmHg, 33 C, sympathetic tone 0.30, recruitment 0.50, oxygen demand 1.0), the model yields a density of 8.0 loops/mm, an erythrocyte velocity of 0.70 mm/s, a capillary SO₂ of 86%, a tissue PO₂ of 36 mmHg, an MFI of 3.0, a heterogeneity index of 21%, an oxygen extraction of 23%, a blood flow of 4.5 mL/100 g/min and a transit time of 0.85 s. These values are internally consistent and lie within the published ranges used as anchors, but they are not calibrated to an individual. The sensitivity analysis in Fig. 6 shows that MAP has the largest effect on flow, with skin temperature and recruitment contributing comparably, while oxygen demand dominates tissue PO₂; the model is intended to expose those relationships, not to predict a patient’s state.

**Fig. 6.**
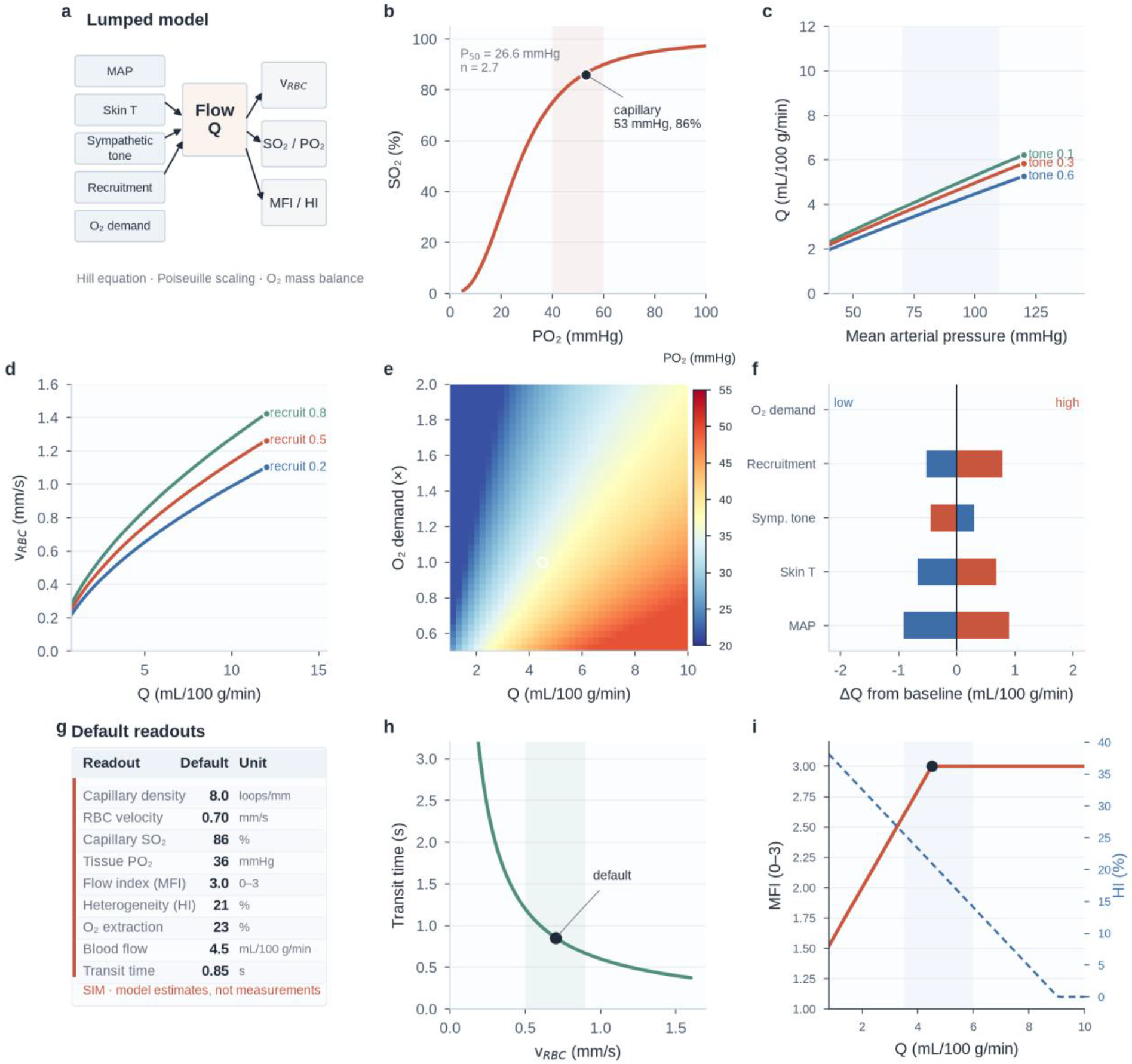
Lumped microcirculatory model and sensitivity. **a**, Model inputs and outputs. **b**, Hill curve with the default capillary operating point (53 mmHg, 86%). **c**, Flow versus mean arterial pressure at three sympathetic tones. **d**, Erythrocyte velocity versus flow at three recruitment levels. **e**, Tissue PO₂ as a function of flow and oxygen demand; the white circle marks the default. **f**, Tornado plot of the change in flow from baseline for low and high parameter values. **g**, Default readouts. **h**, Transit time versus erythrocyte velocity. **i**, MFI and heterogeneity index versus flow. All values are SIM model estimates.

## Contrast and spatial sampling limits of conventional clinical X-ray imaging for hand capillaries

The physics module evaluates whether conventional clinical X-ray imaging can resolve individual capillaries in the living hand. Under the imaging conditions considered, capillary visibility is limited by insufficient attenuation contrast and inadequate spatial sampling (Fig. 7). For unenhanced blood, the primary limitation is the small attenuation difference between blood and surrounding soft tissue. At 60 keV, their mass attenuation coefficients are approximately 0.2053 and 0.2025 cm^2^/g, respectively, corresponding to a difference of approximately 1.4% [9]. With respective densities of 1.06 and 1.03 g/ cm^3^, the linear attenuation coefficients differ by approximately 0.009 cm^−1^, or 4.4%. A 1-mm column of unenhanced blood therefore produces approximately 0.09% transmission contrast, whereas a 10-µm capillary produces approximately 9 × 10^−4^% before pixel averaging. Averaging within a 150-µm pixel reduces the estimated contrast to approximately 6 × 10^−5^%.

**Fig. 7.**
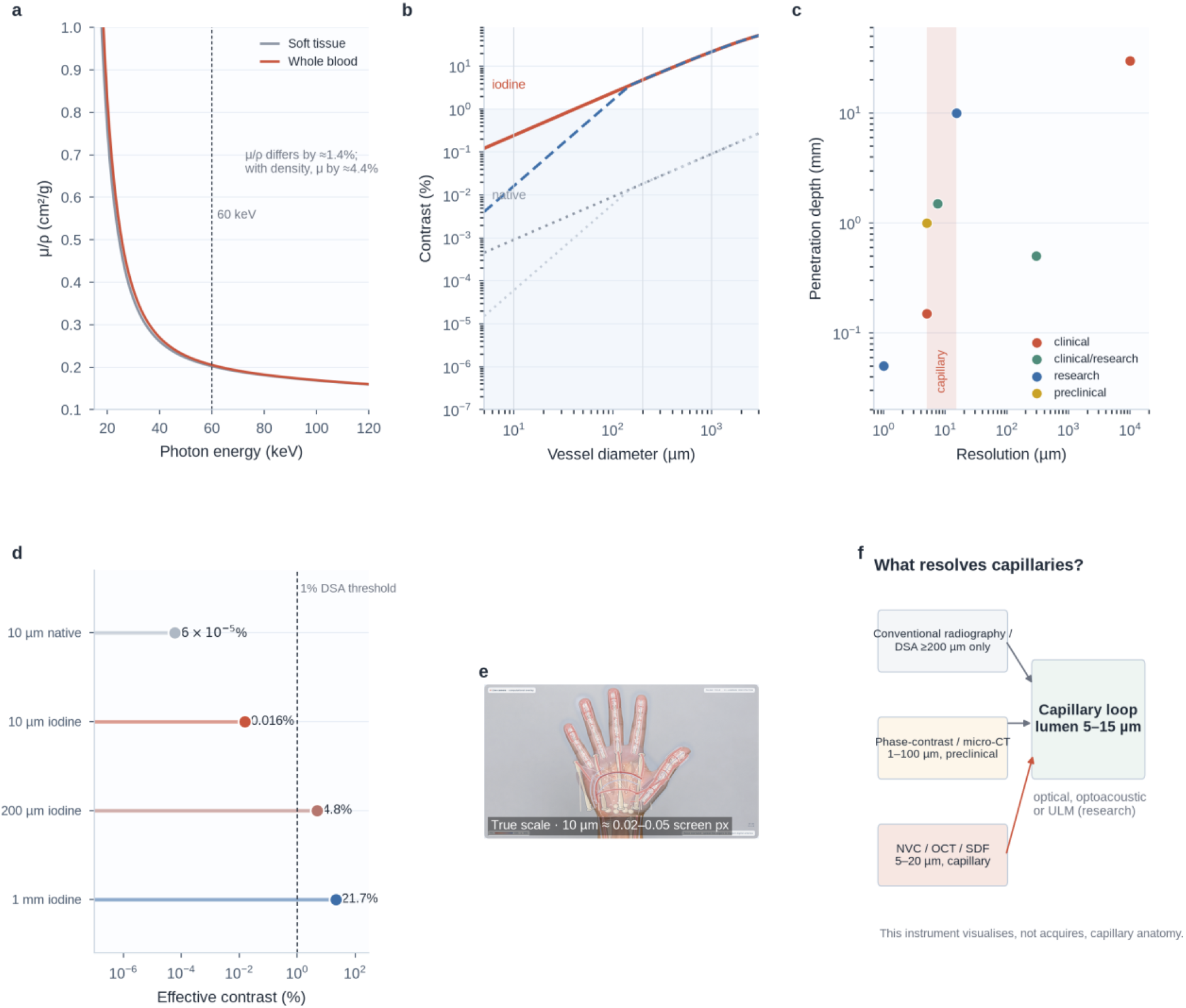
Contrast and spatial sampling limitations of conventional clinical X-ray imaging for individual hand capillaries in vivo. **a**, Mass attenuation coefficients of soft tissue and whole blood from 15 to 120 keV; the dashed line marks 60 keV. **b**, Iodine and native-blood contrast, and the effective contrast in a 150-µm pixel, versus vessel diameter; the capillary, DSA and 1-mm scales are marked. **c**, Resolution–depth map of ten imaging modalities; the shaded band is the capillary lumen. **d**, Effective contrast for a 10-µm native-blood capillary, a 10-µm iodine-filled capillary, a 200-µm arteriole and a 1- mm artery; the dashed line is a 1% DSA detection threshold. **e**, True-scale view; a 10-µm capillary is 0.02–0.05 display px. **f**, Decision summary: optical, optoacoustic and ultrasound localization microscopy can resolve the capillary loop. The calculation is reproducible from the xraylib/NIST attenuation data and the detector sampling range.

Iodinated contrast material at 350 mg I/mL increases the linear attenuation difference to approximately 2.44 cm^−1^, more than two orders of magnitude above that of unenhanced blood. The corresponding transmission contrasts are approximately 21.7% for a 1-mm vessel, 4.8% for a 200-µm vessel, and 0.24% for a 10-µm vessel. After averaging within a 150-µm pixel, the estimated contrast of the 10-µm vessel falls to approximately 0.016%, about 60 times below the representative 1% DSA detection threshold used in the analysis. Contrast enhancement therefore does not overcome the combined effects of small vessel diameter and pixel averaging under these conditions.

Spatial sampling imposes an additional constraint. Clinical digital radiography and computed radiography detectors typically have pixel pitches of 100–200 µm, corresponding to Nyquist frequencies of 2.5–5 line pairs/mm. Sampling a 10-µm vessel across two pixels would require a pixel pitch of no more than 5 µm, corresponding to approximately 100 line pairs/mm and a sampling frequency 20–40 times higher than that of these clinical detectors. DSA can demonstrate arteries approximately 200 µm in diameter [26], but does not resolve individual capillary loops measuring 5–15 µm. Clinical CT generally provides coarser spatial resolution of approximately 0.5–0.7 mm, although ultra-high-resolution photon- counting CT can approach 150 µm. In the modeled CT scenario, partial-volume dilution reduces the signal from a 39-µm loop to well below 1 HU, beneath typical CT noise levels. Even the less stringent criterion of separating adjacent loops remains unmet under the assumed clinical sampling conditions. An inter-capillary spacing of approximately 143 µm requires sampling at intervals of no more than 72 µm, corresponding to approximately 7 line pairs/mm and exceeding the sampling capabilities considered for clinical DR/CR, DSA, and CT.

The module compares radiography, DSA, phase-contrast/micro-CT, NVC, OCT/OCTA, SDF/IDF, laser speckle contrast, NIRS/diffuse optics, photoacoustic microscopy and ultrasound localization microscopy on a resolution–depth map (Fig. 7c). Optical methods (NVC, OCT/OCTA, SDF/IDF) and, in research settings, photoacoustic microscopy and ultrasound localization microscopy can reach the capillary scale in vivo [27]. Research-grade X-ray phase-contrast and micro-CT can reach micrometre voxels in small excised samples [28], but their true resolution is coarser than their voxel size, they are not routine clinical hand imaging, and, to our knowledge, no published study has demonstrated individual capillary loops in a living hand. Conventional radiography, DSA and clinical CT do not resolve 5–15-µm loops in routine hand imaging. The true-scale view illustrates this spatial constraint directly on the hand, where individual capillaries occupy less than one pixel and are labeled as subpixel structures. Together, the resolution–depth comparison and true-scale visualization provide a reproducible quantitative analysis of imaging limitations without relying on patient data or making claims of clinical validation.

## Deployment, performance and the phone

The application is a single-file, client-side system. In camera mode, frames are processed in the browser and are never uploaded; there is no server-side database and no patient data path. For phone use, serve.py –lan –https binds all interfaces, generates a self-signed certificate containing the LAN IPs, prints a phone URL and serves a mobile access page with a QR code, a device preflight (HTTPS, camera, WebGL, touch, installability), a one-minute setup and troubleshooting (Fig. 8). LAN mode generates a one-time token by default; the QR code carries it, and the first visit exchanges it for an HttpOnly cookie. The site is installable as a PWA, with an app-shell cache for offline demonstration.

**Fig. 8.**
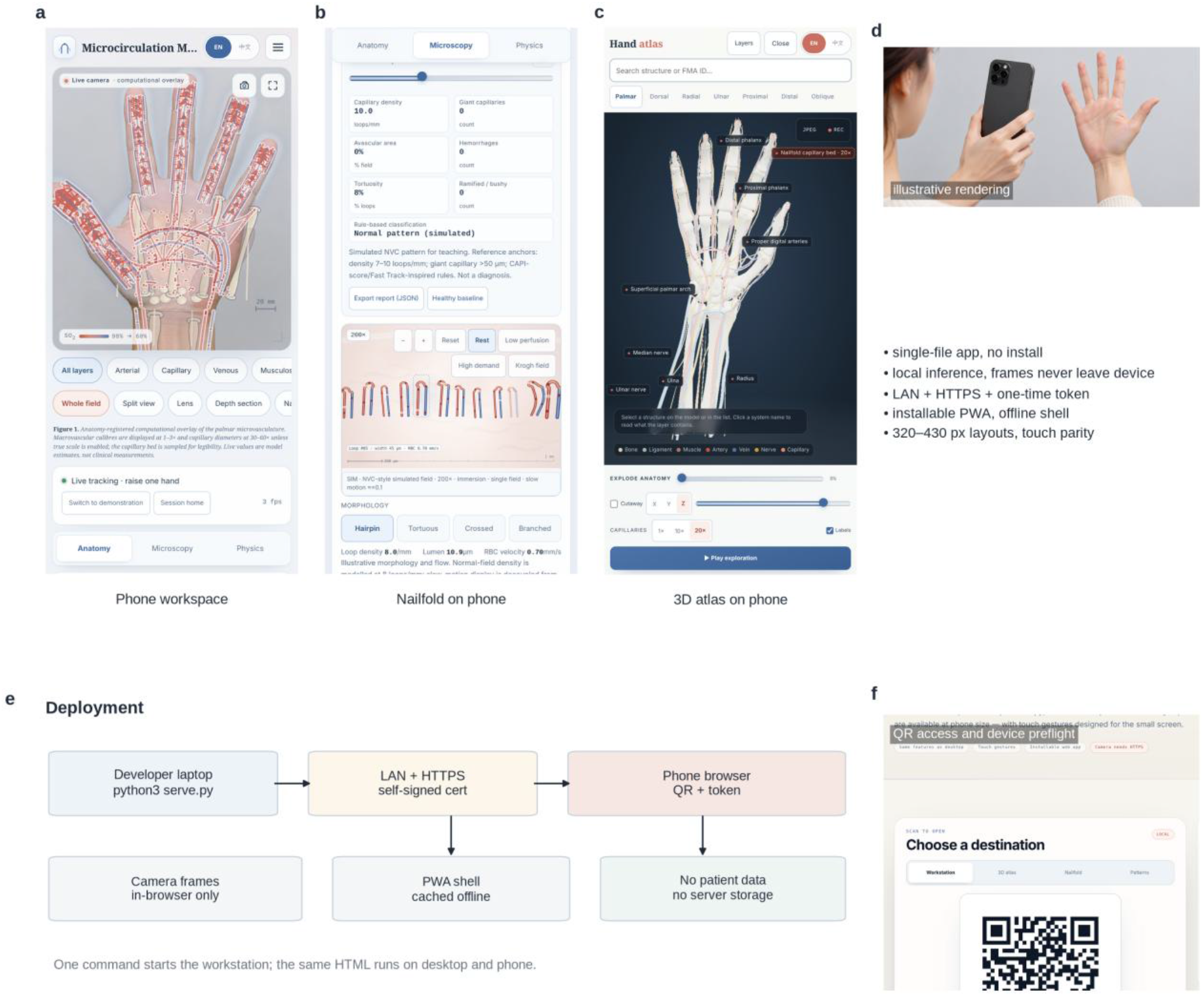
Phone deployment and mobile access. **a**, Phone workspace. **b**, Nailfold microscopy on a phone. **c**, 3D atlas on a phone. **d**, Phone-at-hand use (illustrative rendering) and deployment properties. **e**, Deployment diagram: local server, LAN HTTPS, token, PWA, no patient-data storage. **f**, QR access and device preflight.

The phone layout is the same application, not a separate build. It detects a phone viewport and rearranges the masthead, stage, presets, tabs and controls; the stage can expand to full screen; touch gestures support vessel picking (22 px tolerance), nailfold pinch-zoom and pan, and atlas orbit/pinch; and the camera preview uses an object-fit: cover landmark mapping so the overlay stays registered when the stage is taller than the video frame (Supplementary Fig. 5). Performance controls include device-pixel- ratio caps, low-power blur reduction, off-screen and hidden-tab pauses, microscopy rendering gated to its visible tab, a 960×540 phone video stream, and a merged six-draw-call atlas tier.

On the 2D stage, a frame with about 930 vessel paths plus anatomy renders in 2.9 ms; the nailfold field renders in 1.0 ms; the frame budget at 60 fps is 16.7 ms (Fig. 1d). In the 3D atlas, merging the six anatomical systems into six GPU meshes reduces anatomy draw calls from 136 to 6 (35 total including capillary patches, rings, feeding vessels and helpers), while retaining per-structure selection through invisible pick proxies. Render-on-demand means the atlas does not redraw when the camera and layers are still. On a software rasteriser (SwiftShader), the atlas remains CPU-limited; the performance path reduces draw calls and idle work but does not make software rasterisation equivalent to a hardware GPU. The performance numbers are engineering benchmarks, not clinical performance claims (Supplementary Fig. 3).

## Discussion

*Microcirculation Mirror* integrates multiscale anatomical exploration, representation of evidence provenance, and imaging-physics calculations within a browser-based instrument. These capabilities address a challenge in microvascular visualization, namely the difficulty of relating macroscopic vascular anatomy to microscopic structures while preserving spatial context and communicating the evidential basis of each representation. The principal contribution is therefore not any individual visualization, but an integrated environment in which users can examine anatomical relationships, explore simulated mechanisms, and inspect the limitations of the displayed information.

The instrument supports continuous exploration across anatomical scales. Users can begin with the palmar arches on a tracked hand, follow the digital arteries, visualize the capillary bed, and open a simulated nailfold field. Enabling the true-scale view illustrates how individual capillaries become smaller than a single pixel. A shared hand reference, coordinate framework, and interaction model connect structures spanning nearly three orders of magnitude in vessel diameter. Unlike static illustrations or conventional atlases, the integrated environment maintains spatial and interactive continuity as users move between scales. Pose registration therefore offers a means of linking anatomical levels that are typically presented separately, although its educational value remains to be established empirically.

An equally important feature is the clear identification of information sources and their evidential status. The interface differentiates tracked hand pose from template anatomy, mesh-derived structures from procedural capillaries, published reference values from model estimates, and simulated patterns from patient images. Information about provenance is integral to interpreting the visualizations, not merely supplementary annotation. Labeling bones as templates, the capillary bed as a sampled representation, and the Krogh field as a normalized visual approximation helps prevent visual realism from being mistaken for subject-specific accuracy. Simulation makes selected mechanisms and assumptions available for examination, while source labels clarify that the visualizations do not represent clinical measurements.

The integration of imaging physics extends the instrument beyond anatomical visualization. The limitations of conventional clinical radiography are examined through attenuation coefficients, detector sampling, and true-scale visualization. Under the assumptions used in the module, a 10-µm capillary occupies approximately 0.02–0.05 pixels at true scale, while its effective contrast after pixel averaging is approximately 0.016% when filled with iodinated contrast material; the corresponding contrast for unenhanced blood is approximately 270 times lower. These estimates illustrate how vessel dimensions, attenuation differences, and spatial sampling jointly constrain visibility. They also contextualize the use of nailfold video capillaroscopy, optical coherence tomography, and side stream dark-field imaging for microvascular observation, alongside emerging applications of photoacoustic imaging and ultrasound localization microscopy. The calculations apply to the specified conventional clinical imaging conditions and do not establish a universal limit on X-ray visualization of capillary-scale vessels.

These contributions must be interpreted within the scope of a prototype that has neither incorporated patient data nor undergone clinical validation. The anatomical layers are pose-registered templates, the vascular network is schematic, and the capillary bed is sampled at a density substantially below that of the represented tissue. The nailfold visualization is limited to a single illustrative 1.2-mm field and does not constitute a complete four-field clinical acquisition. The six simulated patterns are generated from literature-informed parameters, with no patient measurements used in their construction. Physiological outputs are derived from a lumped model without individual calibration, and the microvascular flow index and heterogeneity index are empirical approximations. The application is not a medical device and must not be used for diagnosis, screening, risk stratification, or treatment decisions. These limitations informed the interface design, particularly the separation of tracked, template-based, and simulated content.

The present evaluation is restricted to technical characterization, including structural coverage, frame time, draw calls, file size, imaging-physics calculations, and simulator determinism. It does not establish usability, educational benefit, or agreement with a clinical reference standard. Further evaluation could compare learning outcomes or task performance between users of the instrument and users of a conventional atlas. Think-aloud studies could examine how users interpret true-scale views and whether provenance labels support accurate understanding of reference anatomy and simulated content. Expert review of the simulated nailfold patterns could assess their morphological plausibility and consistency with the intended categories, but would not constitute clinical validation of the classification system. Landmark registration should also be evaluated against an independent marker-based reference. Deterministic pattern generation, structured JSON export, and single-file deployment provide a basis for reproducible testing, although these studies have not yet been conducted.

Greater anatomical fidelity represents another direction for development. The current instrument combines open reference meshes for macro-anatomy with procedural representations of microanatomy. Incorporating subject-specific optical coherence tomography or nailfold video capillaroscopy data, or replacing the procedural capillary bed with a micro-CT-derived scaffold, would move the corresponding layer toward an image-derived anatomical model. These extensions would require revised provenance labels and validation procedures appropriate to the acquisition and reconstruction methods. The modular capillary layer and system-level provenance metadata provide an architectural basis for incorporating acquired nailfold fields, segmented optical coherence tomography volumes, and patient-specific meshes while preserving information about the origin and processing of acquired and simulated content.

The deployment approach may also broaden access to computational anatomy. A single HTML file designed to run offline on a laptop or phone, without installation or a patient-data workflow, reduces some of the infrastructural requirements associated with conventional visualization platforms. The documented deployment pathway using a local network, HTTPS, token-based access, and progressive web application functionality is intended to support reproducible demonstrations. It allows the same instrument to be shared with research or clinical teams without requiring a vendor platform, license server, or cloud account. The MIT-licensed code and CC BY-SA atlas further support reuse within their respective licensing conditions. Taken together, these features position *Microcirculation Mirror* as a research and demonstration instrument and as an architectural reference for computational anatomy projects that seek to combine interactive visualization with communication of evidential limits.

## Limitations

*Microcirculation Mirror* remains a research and demonstration prototype with limitations in anatomical sampling, model formulation, and technical implementation. The macroscopic view includes approximately 12 capillary loops per fingertip and 60 per hand, a sparse representation that does not reproduce tissue-level capillary density or network topology. The rule-based categorization of the six simulated nailfold patterns is not a validated implementation of Fast Track or CAPI-score and is intended only for illustration. The visualized Krogh field is normalized and radially exaggerated, with the analytical solution provided separately for comparison; its spatial extent and gradients therefore do not constitute quantitative predictions of tissue oxygenation. The empirical microvascular flow and heterogeneity indices have likewise not been validated against measurement-based indices. Atlas rendering remains constrained by CPU rasterization, and the optimized rendering path does not eliminate the need for GPU acceleration for efficient performance. The application supports tracking of only one hand at a time, while camera-based operation on mobile phones requires WebGL and HTTPS. Single-file deployment simplifies distribution but does not guarantee equivalent functionality or performance across devices and browsers.

## Ethics and Reporting

All relevant ethical guidelines were followed. This study did not involve human participants, patient data, human tissue or identifiable images; therefore, IRB/ethics committee approval and patient/participant consent were not required, and no institutional forms were required or archived. No clinical trial was conducted or registered. The hand images in the figures are illustrative renderings prepared for this article and do not depict a real person.

## Methods

### Software architecture

The application is a single HTML file assembled from modular JavaScript sources by tools/build.py. It uses no runtime framework. The 2D stage is rendered with Canvas2D across four stacked canvases (shade, anatomy, vessels, HUD); the 3D atlas uses Three.js r140 with OrbitControls and GLTFLoader. Hand tracking uses MediaPipe Tasks Vision 0.10.14 [29] with a local float16 hand_landmarker.task model and a CDN fallback. The 3D atlas uses Three.js [30]; MP4 export uses a local H.264 WASM encoder [31]. The project began as a migration of the single-file NeuroMirror viewer [32]. The code is MIT-licensed.

## Hand tracking and coordinate frames

MediaPipe returns 21 landmarks per hand in normalized image coordinates. The application mirrors the x coordinate for the front camera, converts to display coordinates, applies an object-fit: cover mapping when the video aspect differs from the stage, and smooths the landmarks with an exponential filter whose time constant is frame-rate aware. From the smoothed landmarks it builds a palm frame (wrist, index MCP, little MCP), sixteen bone frames (four per finger) and a forearm frame. Each frame is an orthonormal basis scaled by the distance between its defining joints. Detections are considered lost after 18 consecutive frames without a hand; the overlay is then cleared.

## Anatomical templates and atlas extraction

The camera overlay’s anatomy is generated procedurally from canonical hand proportions and the tracked frames. The 3D atlas is extracted from the Z-Anatomy Startup.blend (Blender 3.6.5) with a right- hand region *x* ∈ [−0.38, −0.15], *y* ∈ [−0.12,0.10], *z* ∈ [0.69,1.00] m and a left-hand region mirrored in x. Mesh and curve objects in the Skeletal, Joints, Muscular, Cardiovascular and Nervous collections are evaluated, world transforms are baked, the proximal end is clipped at *z* = 1.00 m, and one GLB is exported per system with one node per structure. Metadata are written to atlas-meta.json; the left-hand set is a separate extraction. The capillary layer is generated at run time from the literature parameters in Supplementary Fig. 2b.

## Nailfold simulator

The NVC field is 1.2 mm wide with 1 mm = 500 px. Loop spacing is 1/*ρ*, where *ρ* is the pattern density in loops/mm; avascular gaps are one to four contiguous intervals whose total width is proportional to the avascular fraction; each loop receives a seeded morphology (hairpin, tortuous, bushy, ramified or giant); giant loops are forced for SSc patterns; haemorrhages are drawn as sub-papillary deposits; and loop width, height and perfusion are modulated by the physiological state. The random seed is deterministic per pattern and index. Metrics are computed from the generated field and reported with the pattern profile, reference ranges and a non-diagnostic disclaimer.

## Physiology model

The model is defined by the equations in the Results and in Supplementary Fig. 2. The Hill parameters are *P*_50_ = 26.6 mmHg and *n* = 2.7 . The Krogh display uses a normalised radial approximation,

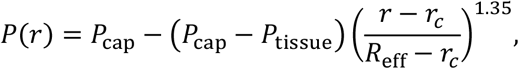

with *r_c_* = 5 µm and *R*_eff_ = 26 µm. For comparison, the steady-state Krogh solution is

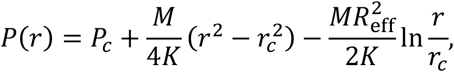

with *K* = 9.4 × 10^−10^ cm^2^ s^−1^ mmHg^−1^ and *M* = 5 × 10^−4^ cm^3^ O₂ cm^−3^ s^−1^ for the resting case. The display field is radially exaggerated and is labelled as: at the default capillary PO₂ of 53 mmHg it drops to 36 mmHg over 26 µm, whereas the physical solution at *M* = 5 × 10^−4^ cm^3^ O₂ cm^−3^ s^−1^ drops only ≈2 mmHg. The 26-µm radius is extrapolated from skeletal-muscle Krogh geometry, not measured in skin.

## Imaging-physics calculations

Mass attenuation coefficients were computed from ICRU-4 soft-tissue and ICRU-44 blood compositions with xraylib 4.x element cross-sections; the iodine value uses the elemental coefficient at 60 keV and a 350 mg I/mL column. Linear attenuation is *μ* = (*μ*/*ρ*)*ρ* with *ρ* = 1.03 g/cm^3^ for soft tissue and 1.06 g/cm^3^ for blood. Projection contrast is *C* = 1 − exp[−(*μ_I_* − *μ_T_*)*d*]; the effective contrast in a detector pixel is *C*min(1, *d*/*p*) for pixel pitch *p*. The detector sampling range (100–200 µm) and the DSA limit (≈200 µm) are taken from the literature [26]. The modality resolution–depth map uses the values in Supplementary Table S2.

## Deployment and security

serve.py is a Python standard-library HTTP server. –lan binds all interfaces; –https generates a self- signed certificate with the LAN IPs in its SAN list; –cert/–key accept an existing certificate; –no-auth disables the one-time token; –token fixes a known value. The server pre-compresses HTML, JS, CSS, JSON, WASM and GLB assets, sets CSP, X-Frame-Options: DENY, COOP/CORP and Permissions- Policy headers, blocks src/, tools/, dotfiles and .py/.pem/.key paths, and disables directory listing. The PWA uses a manifest and a service worker; the app shell is cached for offline demonstration, while / mcm/* endpoints are not cached.

## Evaluation scope

The evaluation is technical and reproducible: structure counts, FMA coverage, file sizes, frame times, draw calls, transfer sizes, the X-ray contrast calculation, the Krogh comparison, and the determinism of the pattern generator. No human participants were involved. No usability, user- performance or clinical accuracy study was performed. The performance measurements were made in Chrome 151 on a Linux host with SwiftShader for the 3D atlas; the 2D frame times are from the application’s own instrumentation and are reported as engineering benchmarks.

## Illustrative hand images

All hand images in the figures are illustrative computational renderings prepared for this article. No human participants were photographed. The images were supplied to the application as a virtual camera stream; the tracker detected 21 landmarks, and the overlays in Figs. 1, 2, 7 and 8 are the application’s own rendering on those images. The illustrative images demonstrate operation and registration.

## Data and code availability

No patient data were generated or analyzed. Source data for all quantitative panels, application source code, the build script, and atlas metadata are available from the corresponding author upon reasonable request. The atlas geometry is derived from Z-Anatomy (CC BY-SA 4.0) and BodyParts3D (CC BY 4.0), and the application code is licensed under the MIT License.

## Author contributions

All authors contributed to the study design, data interpretation and revision of the manuscript. M.L. conceived the instrument, implemented the software, performed the technical evaluation, generated the figures and wrote the first draft. All authors read and approved the final manuscript.

## Data Availability

All data produced in the present study are available upon reasonable request to the authors

## Acknowledgements

The 3D atlas derives from Z-Anatomy and BodyParts3D (Database Center for Life Science, Japan). The application uses MediaPipe Tasks Vision, Three.js and h264-mp4-encoder. The author thanks the open-anatomy community for maintaining the source meshes.

