## Supplementary Information for "Microcirculation Mirror: A zero-install browser workstation for hand microvascular anatomy and nailfold capillaroscopy"

**Running title: Browser-based microvascular anatomy workstation**

Yingyu Wang[1,2], Jingjing He[1,2], Weiqi Liu[1,2], Xiang Xu[1,2], Bingwei Li[1,2], Yuan Li[1,2], Bing Wang[1,2], Mingming Liu[1,2,*]

[1]Institute of Microcirculation, Chinese Academy of Medical Sciences & Peking Union Medical College, Beijing 100005, China

[2]International Center of Microvascular Medicine, Chinese Academy of Medical Sciences, Beijing 100005, China

[*]Correspondence: Mingming Liu, Institute of Microcirculation, Chinese Academy of Medical Sciences & Peking Union Medical College, No.5 Dong Dan Third Alley, Dongcheng District, Beijing 100005, China..

#### Supplementary Methods

##### Software package and build

The application is assembled from modular JavaScript sources by tools/build.py; the build in lines CSS and JavaScript into a single index.html. The optional local assets are a MediaPipe Tasks Vision 0.10.14 runtime, a float16 hand-landmark model (7.5 MB), six GLB layers per hand and a Three.js r140 build. The 2D stage uses Canvas2D; the 3D atlas uses WebGL. No server-side database or user account is used.

##### Atlas extraction and validation

The Z-Anatomy Startup.blend was loaded headless in Blender 3.6.5. Right- and left-hand regions were defined in Z-Anatomy coordinates; mesh and curve objects in the Skeletal, Joints, Muscular, Cardiovascular and Nervous collections were evaluated, world transforms baked, the proximal end clipped at $z = 1.00$ m, and one GLB exported per system with one node per structure. Metadata (English and Chinese names, FMA identifiers, descriptions, bounding boxes, vertex and polygon counts) were written to atlas-meta.json. The shipped model contains 136 structures per hand; 36 carry an FMA identifier. The atlas was checked with tools/validate_atlas.py for missing names, empty meshes and out-of-range bounds.

##### Nailfold pattern generator

The field is 1.2 mm wide with 1 mm = 500 px. Loop spacing is $1/\rho$; avascular gaps are one to four contiguous intervals whose total width is proportional to the avascular fraction; each loop receives a seeded morphology; giant loops are forced for SSc patterns; haemorrhages are drawn as sub-papillary deposits. The random seed is deterministic per pattern and index. The six profiles and their nominal severities are listed in Supplementary Table S4; the table gives profile values at severity = 1.0, while Fig. 5g and the interface show the values blended toward the healthy baseline at the nominal severity, $m(s) = m_h + s(m_p - m_h)$. The severity slider blends each profile with the healthy baseline, $m(s) = m_h + s(m_p - m_h)$. Metrics are computed from the generated field.

#### Physiology model

The flow index, blood flow, erythrocyte velocity, Hill saturation, oxygen extraction, venous saturation, capillary saturation, tissue $PO_2$, MFI, heterogeneity index and transit time are computed as described in the main Methods. The display Krogh field is a normalized approximation; the physical Krogh solution is shown for comparison with $K = 9.4 \times 10^{-10}$ cm$^2$ s$^{-1}$ mmHg$^{-1}$ and $M = 5 \times 10^{-4}$ cm$^3$ $O_2$ cm$^{-3}$ s$^{-1}$.

#### Imaging-physics calculation

Mass attenuation coefficients were computed from ICRU-4 soft-tissue and ICRU-44 blood compositions using xraylib element cross-sections. Linear attenuation used $\rho = 1.03$ g/cm$^3$ (soft tissue) and 1.06 g/cm$^3$ (blood). Iodine contrast used the elemental coefficient at 60 keV and a 350 mg I/mL column. Projection contrast is $C = 1 - \exp[-(\mu_I - \mu_T)d]$; effective contrast in a pixel is $C\min(1, d/p)$.

#### Reproduction

The screenshots were captured with Chrome 151 through the DevTools Protocol; the illustrative palm was supplied as a virtual camera stream.

### Supplementary Figures

#### Supplementary Fig. 1 | Complete structure inventory of the hand atlas

136 named structures per hand, grouped by system. FMA identifiers are available for 36 structures; all structures carry English and Chinese names. Vertex and polygon counts are from the shipped GLB meshes.

| System | Structure | 中文名 | FMA | Verts |
|---|---|---|---|---|
| | Capitate bone | 头状骨 | FMA24446 | 238 |
| | Distal phalanx of fifth finger of hand | 小指远节指骨 | FMA23949 | 98 |
| | Distal phalanx of first finger of hand | 拇指远节指骨 | FMA23945 | 113 |
| | Distal phalanx of fourth finger of hand | 环指远节指骨 | FMA23948 | 69 |
| | Distal phalanx of second finger of hand | 食指远节指骨 | FMA23946 | 127 |
| | Distal phalanx of third finger of hand | 中指远节指骨 | FMA23947 | 98 |
| | Fifth metacarpal bone | 第五掌骨 | FMA23903 | 258 |
| | First metacarpal bone | 第一掌骨 | FMA23899 | 343 |
| | Fourth metacarpal bone | 第四掌骨 | FMA23902 | 323 |
| | Hamate bone | 钩骨 | FMA24448 | 270 |
| | Lunate bone | 月骨 | FMA24437 | 1046 |
| | Middle phalanx of fifth finger of hand | 小指中节指骨 | FMA23936 | 148 |
| | Middle phalanx of fourth finger of hand | 环指中节指骨 | FMA23935 | 225 |
| | Middle phalanx of second finger of hand | 食指中节指骨 | FMA23933 | 159 |
| | Middle phalanx of third finger of hand | 中指中节指骨 | FMA23934 | 182 |
| | Pisiform bone | 豌豆骨 | FMA24441 | 836 |
| | Proximal phalanx of fifth finger of hand | 小指近节指骨 | FMA23922 | 282 |
| | Proximal phalanx of first finger of hand | 拇指近节指骨 | FMA23918 | 166 |
| | Proximal phalanx of fourth finger of... | 环指近节指骨 | FMA23921 | 186 |
| | Proximal phalanx of second finger of... | 食指近节指骨 | FMA23919 | 209 |
| | Proximal phalanx of third finger of hand | 中指近节指骨 | FMA23920 | 222 |
| | Radius | 桡骨 | FMA23464 | 254 |
| | Scaphoid bone | 手舟骨 | FMA24435 | 197 |
| | Second metacarpal bone | 第二掌骨 | FMA23900 | 384 |
| | Third metacarpal bone | 第三掌骨 | FMA23901 | 356 |
| | Trapezium bone | 大多角骨 | FMA24443 | 252 |
| | Trapezoid bone | 小多角骨 | FMA23725 | 184 |
| | Triquetrum bone | 三角骨 | FMA24439 | 90 |
| | Ulna | 尺骨 | FMA23467 | 164 |
| | Articular capsule of radiocarpal joint | 桡腕关节囊 | — | 759 |
| | Articular capsules of distal... | 远侧指间关节囊 | — | 846 |
| | Articular capsules of... | 掌指关节囊 | — | 880 |
| | Articular capsules of proximal... | 近侧指间关节囊 | — | 778 |
| | Collateral interphalangeal ligaments of... | 指间关节侧副韧带 | — | 1020 |
| | Collateral metacarpophalangeal ligaments | 掌指关节侧副韧带 | — | 540 |
| | Dorsal carpometacarpal ligaments | 腕掌关节背侧韧带 | — | 86 |
| | Dorsal intercarpal ligaments | 腕骨间背侧韧带 | — | 222 |
| | Dorsal radiocarpal ligament | 桡腕背侧韧带 | — | 270 |
| | Interosseous membrane of forearm | 前臂骨间膜 | — | 1170 |
| | Palmar carpometacarpal ligaments | 腕掌关节掌侧韧带 | — | 220 |
| | Palmar interphalangeal ligaments | 指间关节掌侧韧带 | — | 450 |
| | Pisotriquetral ligament | 豌豆三角骨韧带 | — | 222 |
| | Radiate carpal ligament | 腕辐状韧带 | — | 206 |
| | Abductor digiti minimi of hand | 小指展肌 | — | 336 |
| | Abductor pollicis brevis | 拇短展肌 | — | 312 |
| | Abductor pollicis longus | 拇长展肌 | — | 1123 |
| | Antebrachial fascia | 前臂筋膜 | — | 1019 |
| | Brachioradialis muscle | 肱桡肌 | — | 1177 |
| | Common flexor tendon sheath | 屈肌腱总腱鞘 | — | 7094 |
| | Cruciform part of fibrous sheath of... | 手指纤维鞘交叉部 | — | 2382 |
| | Deep head of flexor pollicis brevis | 拇短屈肌深头 | — | 371 |
| | Deep head of pronator teres | 旋前圆肌深头 | — | 105 |
| | Dorsal fascia of hand | 手背筋膜 | — | 819 |
| | Dorsal interossei muscles of hand | 骨间背侧肌 | — | 693 |
| | Extensor carpi radialis brevis | 桡侧腕短伸肌 | — | 992 |
| | Extensor carpi radialis longus | 桡侧腕长伸肌 | — | 751 |
| | Extensor digiti minimi | 小指伸肌 | — | 799 |
| | Extensor digitorum | 指伸肌 | — | 2694 |
| | Extensor indicis | 示指伸肌 | — | 573 |
| | Extensor pollicis brevis | 拇短伸肌 | — | 590 |
| | Extensor pollicis longus | 拇长伸肌 | — | 803 |
| | Extensor retinaculum of wrist | 腕伸肌支持带 | — | 262 |
| | Flexor carpi radialis | 桡侧腕屈肌 | — | 511 |
| | Flexor digiti minimi of hand | 小指短屈肌 | — | 156 |
| | Flexor digitorum profundus | 指深屈肌 | — | 3081 |
| | Flexor pollicis longus | 拇长屈肌 | — | 822 |
| | Flexor retinaculum of wrist | 屈肌支持带 | — | 358 |
| | Humeral head of extensor carpi ulnaris | 尺侧腕伸肌肱头 | — | 304 |
| | Humeral head of flexor carpi ulnaris | 尺侧腕屈肌肱头 | — | 1164 |
| | Humero-ulnar head of flexor digitorum... | 指浅屈肌肱尺头 | — | 4499 |
| | Lumbrical muscles of hand | 蚓状肌 | — | 692 |
| | Oblique head of adductor pollicis | 拇收肌斜头 | — | 227 |
| | Opponens digiti minimi muscle of hand | 小指对掌肌 | — | 210 |
| | Opponens pollicis muscle | 拇对掌肌 | — | 194 |
| | Palmar aponeurosis | 掌腱膜 | — | 749 |
| | Palmar interossei muscles | 骨间掌侧肌 | — | 87 |
| | Palmaris longus muscle | 掌长肌 | — | 1618 |
| | Pronator quadratus | 旋前方肌 | — | 527 |
| | Radial head of flexor digitorum... | 指浅屈肌桡头 | — | 510 |
| | Superficial head of flexor pollicis... | 拇短屈肌浅头 | — | 136 |
| | Superficial head of pronator teres | 旋前圆肌浅头 | — | 258 |
| | Superficial transverse metacarpal... | 掌浅横韧带 | — | 162 |
| | Supinator | 旋后肌 | — | 86 |
| | Synovial sheaths of digits of hand | 手指滑膜鞘 | — | 3494 |
| | Tendon sheath - abd. pollicis longus -... | ... | — | 342 |
| | Tendon sheath of extensor digitorum and... | ... | — | 1186 |
| | Tendon sheath of extensors carpi... | 桡侧腕伸肌腱鞘 | — | 1154 |
| | Transverse head of adductor pollicis | 拇收肌横头 | — | 509 |
| | Ulnar head of extensor carpi ulnaris | 尺侧腕伸肌尺头 | — | 255 |
| | Ulnar head of flexor carpi ulnaris | 尺侧腕屈肌尺头 | — | 453 |
| | Common palmar digital arteries | 指掌侧总动脉 | — | 1632 |
| | Deep palmar arch | 掌深弓 | FMA22839 | 1164 |
| | Dorsal carpal anastomosis | 腕背动脉吻合 | — | 300 |
| | Dorsal carpal branch of ulnar artery | 尺动脉腕背支 | — | 876 |
| | Dorsal digital arteries of hand | 手指背动脉 | — | 7584 |
| | Dorsal metacarpal arteries | 掌背动脉 | — | 2940 |
| | Palmar carpal branch of radial artery | 桡动脉腕掌支 | — | 300 |
| | Palmar metacarpal arteries | 掌心动脉 | — | 2208 |
| | Posterior interosseous artery | 骨间后动脉 | — | 420 |
| | Proper palmar digital arteries | 指掌侧固有动脉 | — | 15672 |
| | Radial artery | 桡动脉 | FMA22733 | 1176 |
| | Superficial palmar arch | 掌浅弓 | FMA22835 | 1452 |
| | Ulnar artery | 尺动脉 | FMA22797 | 504 |
| | Basilic vein | 贵要静脉 | FMA22909 | 731 |
| | Cephalic vein | 头静脉 | FMA13325 | 1079 |
| | Deep venous palmar arch | 掌深静脉弓 | — | 2460 |
| | Dorsal digital veins of hand | 手指背静脉 | — | 9324 |
| | Dorsal venous network of hand | 手背静脉网 | — | 1308 |
| | Median antebrachial vein | 前臂正中静脉 | FMA22968 | 1865 |
| | Palmar digital veins | 指掌侧静脉 | — | 14412 |
| | Radial veins | 桡静脉 | — | 744 |
| | Superficial venous palmar arch | 掌浅静脉弓 | — | 2172 |
| | Ulnar veins | 尺静脉 | — | 936 |
| | (Communicating branch of median nerve... | ... | — | 300 |
| | Anterior branch of medial antebrachial... | 前臂内侧皮神经前支 | — | 180 |
| | Anterior interosseous nerve of forearm | 前臂骨间前神经 | — | 480 |
| | Common palmar digital branches of... | 正中神经掌侧总指支 | — | 1188 |
| | Common palmar digital branches of ulnar... | 尺神经掌侧总指支 | — | 900 |
| | Deep branch of radial nerve | 桡神经深支 | — | 492 |
| | Deep branch of ulnar nerve | 尺神经深支 | — | 732 |
| | Dorsal branch of ulnar nerve | 尺神经手背支 | — | 744 |
| | Dorsal digital branches of radial nerve | 桡神经指背支 | — | 6852 |
| | Dorsal digital branches of ulnar nerve | 尺神经指背支 | — | 2484 |
| | Lateral antebrachial cutaneous nerve | 前臂外侧皮神经 | — | 324 |
| | Median nerve | 正中神经 | — | 576 |
| | Muscular branches of ulnar nerve | 尺神经肌支 | — | 156 |
| | Palmar branch of median nerve | 正中神经掌支 | — | 588 |
| | Palmar branch of ulnar nerve | 尺神经掌支 | — | 588 |
| | Posterior antebrachial cutaneous nerve | 前臂后皮神经 | — | 407 |
| | Posterior branch of medial antebrachial... | 前臂内侧皮神经后支 | — | 264 |
| | Posterior interosseous nerve of forearm | 前臂骨间后神经 | — | 420 |
| | Proper palmar digital branches of... | ... | — | 5556 |
| | Proper palmar digital branches of ulnar... | 尺神经掌侧固有指支 | — | 2916 |
| | Superficial branch of radial nerve | 桡神经浅支 | — | 1776 |
| | Superficial branch of ulnar nerve | 尺神经浅支 | — | 444 |
| | Ulnar nerve | 尺神经 | — | 600 |

Skeleton (29) Ligaments & joints (14) Muscles & sheaths (47)
Arteries (13) Veins (10) Nerves (23)

**Supplementary Fig. 1 Complete structure inventory of the hand atlas.** 136 named structures per hand, grouped by system, with English and Chinese names, FMA identifiers where available, and vertex/polygon counts. The full table is also provided as Supplementary Table S1.

**a**

##### Model equations

Hill saturation

$$S(P) = P^n/(P^n + P_{50}^n)$$

P50 = 26.6 mmHg, n = 2.7

Lumped flow

$$Q = 6.4\,(MAP/90)^{0.9}(1 + 0.05(T - 33))(1 - 0.30S_{sym})(0.55 + 0.45R)$$

clamped to 0.8–12

Erythrocyte velocity

$$v = 0.85\,(Q/4.5)^{0.60}(0.65 + 0.35R)$$

clamped to 0.08–1.8 mm/s

$O_2$ extraction

$$O_2ER = 0.23\,VO_2(4.5/Q)$$

clamped to 8–72%

Krogh cylinder

$$P(r) = P_c + \frac{M}{4K}(r^2 - r_c^2) - \frac{MR^2}{2K}\ln\frac{r}{r_c}$$

r_c = 5 µm, R = 26 µm

Projection contrast

$$C = 1 - \exp[-(\mu_I - \mu_T)d]$$

60 keV, 350 mg I/mL

Flow index (empirical)

$$MFI = 3(0.40 + 0.60\,Q/4.5)$$

0–3 scale

**b**

##### Parameters and literature anchors

| Quantity | Value used | Source | Conf. |
|---|---|---|---|
| Superficial palmar arch | 3.0 ± 0.7 mm | [14] | high |
| Proper digital artery | 0.9 / 0.8 mm | [18] | high |
| Nailfold density | 7–10 loops/mm | [26,27] | high |
| Loop length | 198–215 µm | [28,29] | high |
| Loop outer width | ~39 µm | [28] | medium |
| Limb diameter | 5.6 / 8.7 µm | [24] | high |
| Apex diameter | 13.4 ± 1.5 µm | [24] | high |
| Arterial limb RBC | 0.84 ± 0.53 mm/s | [31] | medium |
| Venous limb RBC | 0.47 ± 0.29 mm/s | [31] | medium |
| Tissue $PO_2$ | 30–40 mmHg | [38] | medium |
| $O_2$ extraction | ~25% (model 23%) | [36] | medium |
| Hill P50 / n | 26.6 mmHg / 2.7 | [37] | high |
| Krogh K | $9.4 \times 10^{-10}$ $cm^2\,s^{-1}\,mmHg^{-1}$ | [35] | medium |
| Krogh R_min | 26 µm (extrapolated) | [35] | low |
| Blood–tissue contrast | 1.4% mass / 4.4% linear at 60 keV | [1,2] | high |
| DR/CR sampling | 100–200 µm | [6] | high |
| DSA arterial limit | ~200 µm | [8] | high |
| NVC resolution | 2.2 µm/px, 1.7 × 1.3 mm | [26,34] | high |

**Supplementary Fig. 2 Model equations and parameter anchors.** The Hill, flow, velocity, oxygen-extraction, Krogh and projection-contrast equations, with the literature values and confidence notes used in the instrument.

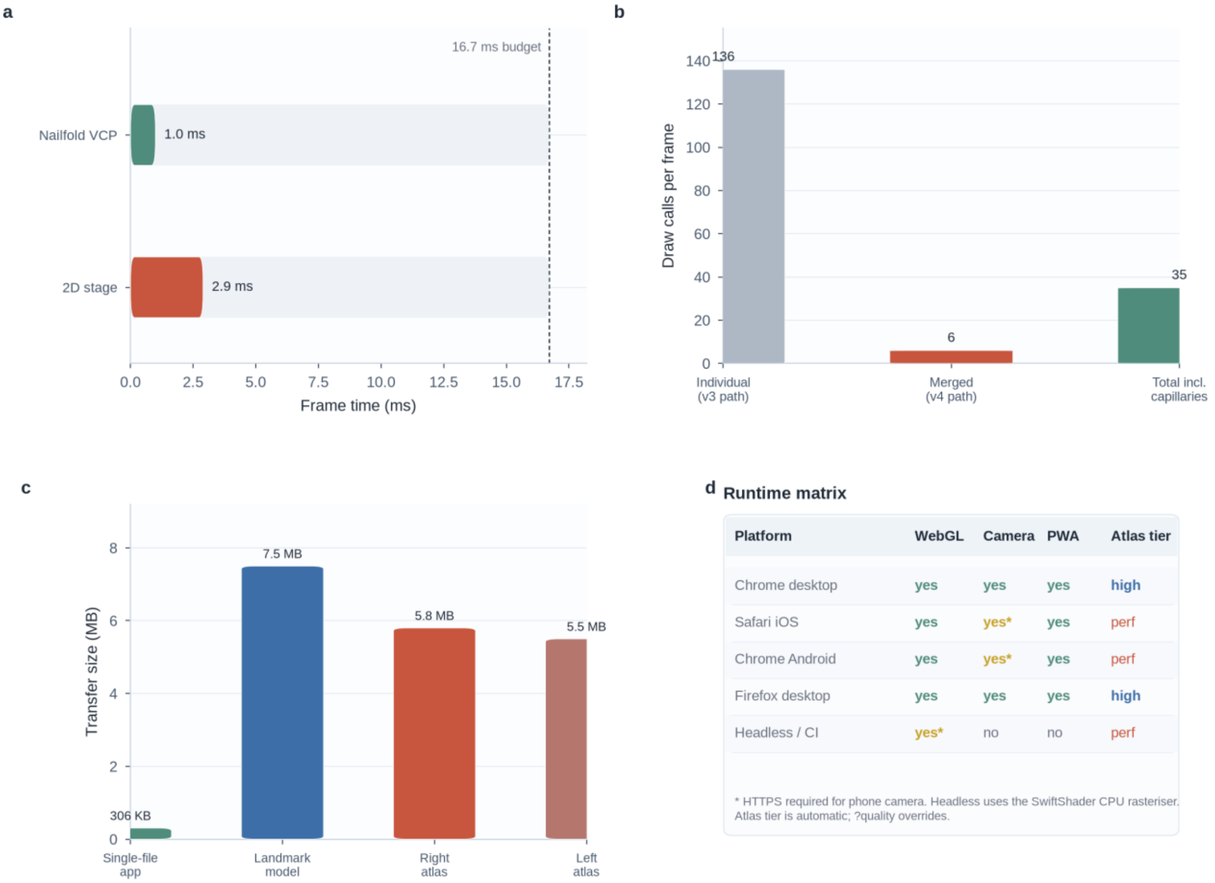


**Supplementary Fig. 3 Technical performance.** Frame budget, draw-call reduction, transfer sizes and the runtime matrix. These are engineering benchmarks, not clinical performance claims.

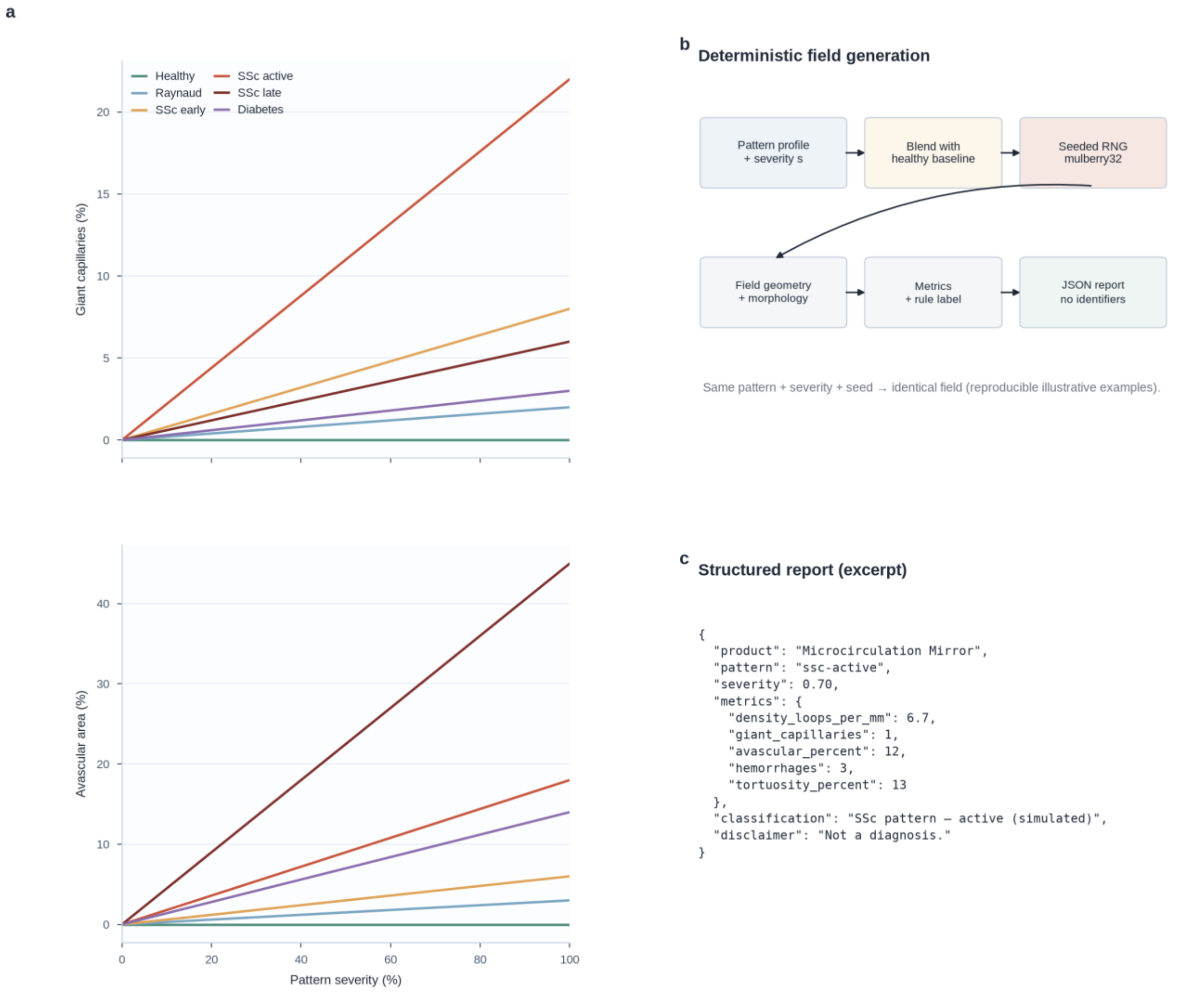


**Supplementary Fig. 4 Deterministic pattern generation.** Severity-blend curves for giant capillaries and avascular width fraction, the generation pipeline, and an excerpt of the structured JSON report.

a b c

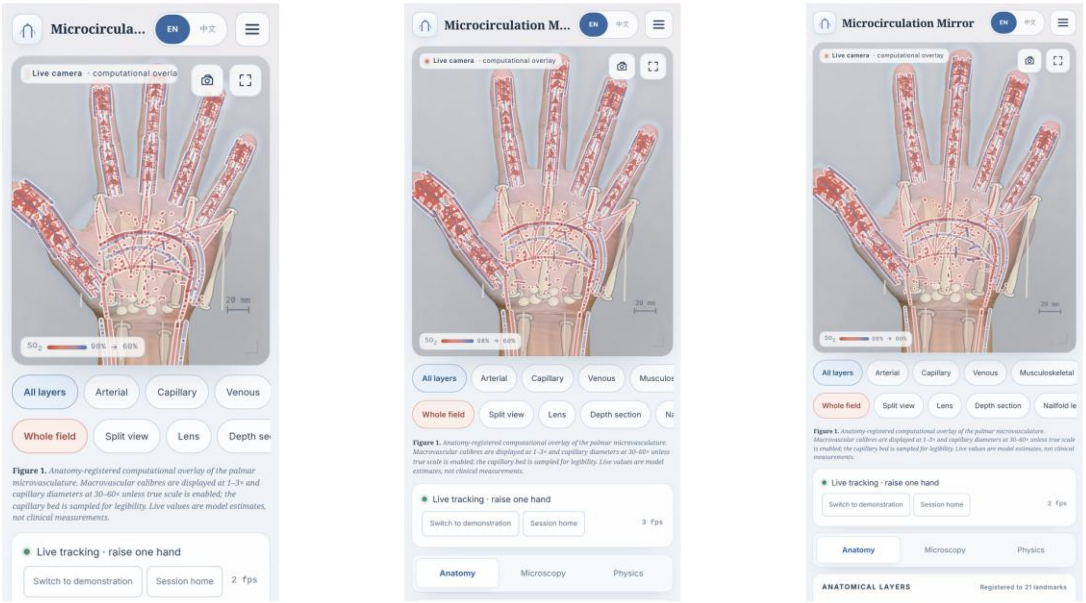


320 px  390 px  430 px

**d** Layout and accessibility audit

| Check | 320 px | 375 px | 390 px | 430 px | Tablet / desktop |
|---|---|---|---|---|---|
| Horizontal overflow | none | none | none | none | none |
| Masthead height | 52 px | 52 px | 52 px | 52 px | 64 px |
| Stage height | ~52% | ~52% | ~52% | ~52% | adaptive |
| Touch target (min) | 44 px | 44 px | 44 px | 44 px | *n/a* |
| Vessel hit tolerance | 22 px | 22 px | 22 px | 22 px | 12 px |
| Camera (HTTPS) | yes | yes | yes | yes | yes |
| PWA installable | yes | yes | yes | yes | yes |

Measured in Chrome 151 at device-pixel ratios 2–3; touch targets follow the 44 px design minimum. No patient data are stored.

**Supplementary Fig. 5 Phone layouts and accessibility audit.** The same application at 320, 390 and 430 px, with the layout and touch-target audit.

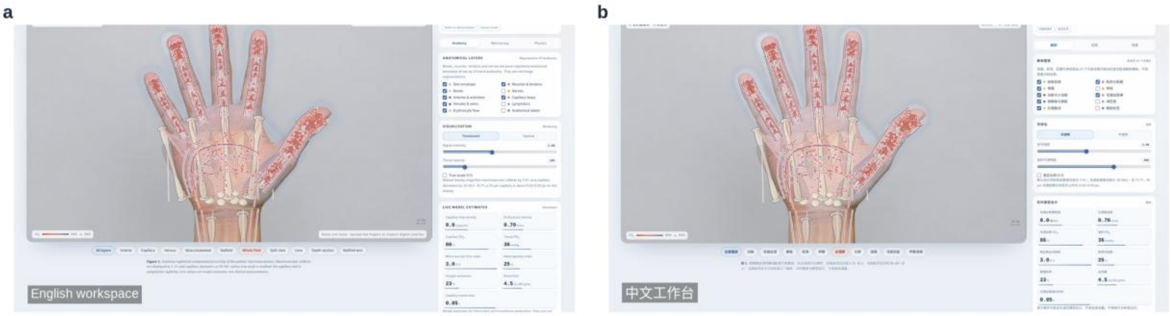


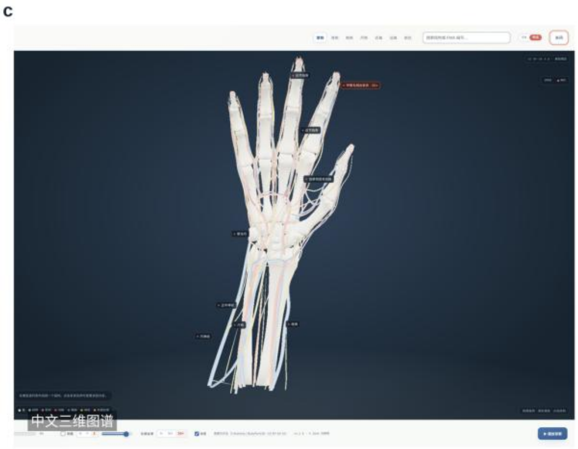


d

**Bilingual coverage**

| Surface | EN | 中文 | Notes |
|---|---|---|---|
| Workspace & layers | 100% | 100% | UI strings |
| Nailfold & patterns | 100% | 100% | metrics, labels |
| 3D atlas structures | 136 | 136 | English + ZH |
| Atlas systems | 6 | 6 | names and blurbs |
| Legal & privacy | 100% | 100% | full text |
| JSON report | EN keys | 中文 names | no identifiers |

Language choice persists in localStorage; anatomical names follow Terminologia Anatomica English equivalents.

**Supplementary Fig. 6 Bilingual interface.** English and Chinese workspaces, the Chinese 3D atlas, and the bilingual coverage audit.

**Supplementary Tables**

**Supplementary Table S1 Complete structure inventory (136 structures per hand).**

| System | Structure | FMA | Vertices | Polygons |
|---|---|---|---|---|
| Skeleton | Capitate bone | FMA24446 | 238 | 472 |
| Skeleton | Distal phalanx of fifth finger of hand | FMA23949 | 98 | 192 |
| Skeleton | Distal phalanx of first finger of hand | FMA23945 | 113 | 222 |
| Skeleton | Distal phalanx of fourth finger of hand | FMA23948 | 69 | 134 |
| Skeleton | Distal phalanx of second finger of hand | FMA23946 | 127 | 250 |
| Skeleton | Distal phalanx of third finger of hand | FMA23947 | 98 | 192 |
| Skeleton | Fifth metacarpal bone | FMA23903 | 258 | 512 |
| Skeleton | First metacarpal bone | FMA23899 | 343 | 682 |
| Skeleton | Fourth metacarpal bone | FMA23902 | 323 | 642 |
| Skeleton | Hamate bone | FMA24448 | 270 | 536 |
| Skeleton | Lunate bone | FMA24437 | 1046 | 2088 |
| Skeleton | Middle phalanx of fifth finger of hand | FMA23936 | 148 | 292 |
| Skeleton | Middle phalanx of fourth finger of hand | FMA23935 | 225 | 446 |
| Skeleton | Middle phalanx of second finger of hand | FMA23933 | 159 | 314 |
| Skeleton | Middle phalanx of third finger of hand | FMA23934 | 182 | 360 |
| Skeleton | Pisiform bone | FMA24441 | 836 | 1668 |
| Skeleton | Proximal phalanx of fifth finger of hand | FMA23922 | 282 | 560 |
| Skeleton | Proximal phalanx of first finger of hand | FMA23918 | 166 | 328 |
| Skeleton | Proximal phalanx of fourth finger of hand | FMA23921 | 186 | 368 |
| Skeleton | Proximal phalanx of second finger of hand | FMA23919 | 209 | 414 |
| Skeleton | Proximal phalanx of third finger of hand | FMA23920 | 222 | 440 |
| Skeleton | Radius | FMA23464 | 254 | 504 |
| Skeleton | Scaphoid bone | FMA24435 | 197 | 390 |
| Skeleton | Second metacarpal bone | FMA23900 | 384 | 764 |
| Skeleton | Third metacarpal bone | FMA23901 | 356 | 708 |
| Skeleton | Trapezium bone | FMA24443 | 252 | 500 |
| Skeleton | Trapezoid bone | FMA23725 | 184 | 364 |
| Skeleton | Triquetrum bone | FMA24439 | 90 | 176 |
| Skeleton | Ulna | FMA23467 | 164 | 324 |
| Ligaments & joints | Articular capsule of radiocarpal joint | – | 759 | 1342 |
| Ligaments & joints | Articular capsules of distal interphalangeal joints | – | 846 | 1504 |
| Ligaments & joints | Articular capsules of metacarpophalangeal joints | – | 880 | 1568 |
| Ligaments & joints | Articular capsules of proximal interphalangeal joints | – | 778 | 1392 |
| Ligaments & | Collateral interphalangeal ligaments of hand | – | 1020 | 1968 |

| | | | | |
|---|---|---|---|---|
| joints | | | | |
| Ligaments & joints | Collateral metacarpophalangeal ligaments | – | 540 | 1040 |
| Ligaments & joints | Dorsal carpometacarpal ligaments | – | 86 | 148 |
| Ligaments & joints | Dorsal intercarpal ligaments | – | 222 | 440 |
| Ligaments & joints | Dorsal radiocarpal ligament | – | 270 | 248 |
| Ligaments & joints | Interosseous membrane of forearm | – | 1170 | 2336 |
| Ligaments & joints | Palmar carpometacarpal ligaments | – | 220 | 416 |
| Ligaments & joints | Palmar interphalangeal ligaments | – | 450 | 864 |
| Ligaments & joints | Pisotriquetral ligament | – | 222 | 264 |
| Ligaments & joints | Radiate carpal ligament | – | 206 | 408 |
| Muscles & sheaths | Abductor digiti minimi of hand | – | 336 | 672 |
| Muscles & sheaths | Abductor pollicis brevis | – | 312 | 624 |
| Muscles & sheaths | Abductor pollicis longus | – | 1123 | 2242 |
| Muscles & sheaths | Antebrachial fascia | – | 1019 | 2004 |
| Muscles & sheaths | Brachioradialis muscle | – | 1177 | 2350 |
| Muscles & sheaths | Common flexor tendon sheath | – | 7094 | 13968 |
| Muscles & sheaths | Cruciform part of fibrous sheath of digit of hand | – | 2382 | 4728 |
| Muscles & sheaths | Deep head of flexor pollicis brevis | – | 371 | 746 |
| Muscles & sheaths | Deep head of pronator teres | – | 105 | 206 |
| Muscles & sheaths | Dorsal fascia of hand | – | 819 | 1445 |
| Muscles & sheaths | Dorsal interossei muscles of hand | – | 693 | 1364 |
| Muscles & | Extensor carpi radialis brevis | – | 992 | 1980 |

| | | | | |
|---|---|---|---|---|
| sheaths | | | | |
| Muscles & sheaths | Extensor carpi radialis longus | – | 751 | 1493 |
| Muscles & sheaths | Extensor digiti minimi | – | 799 | 1594 |
| Muscles & sheaths | Extensor digitorum | – | 2694 | 5396 |
| Muscles & sheaths | Extensor indicis | – | 573 | 1142 |
| Muscles & sheaths | Extensor pollicis brevis | – | 590 | 1176 |
| Muscles & sheaths | Extensor pollicis longus | – | 803 | 1602 |
| Muscles & sheaths | Extensor retinaculum of wrist | – | 262 | 368 |
| Muscles & sheaths | Flexor carpi radialis | – | 511 | 1018 |
| Muscles & sheaths | Flexor digiti minimi of hand | – | 156 | 308 |
| Muscles & sheaths | Flexor digitorum profundus | – | 3081 | 6160 |
| Muscles & sheaths | Flexor pollicis longus | – | 822 | 1644 |
| Muscles & sheaths | Flexor retinaculum of wrist | – | 358 | 524 |
| Muscles & sheaths | Humeral head of extensor carpi ulnaris | – | 304 | 616 |
| Muscles & sheaths | Humeral head of flexor carpi ulnaris | – | 1164 | 2324 |
| Muscles & sheaths | Humero-ulnar head of flexor digitorum superficialis | – | 4499 | 8994 |
| Muscles & sheaths | Lumbrical muscles of hand | – | 692 | 1368 |
| Muscles & sheaths | Oblique head of adductor pollicis | – | 227 | 449 |
| Muscles & sheaths | Opponens digiti minimi muscle of hand | – | 210 | 420 |
| Muscles & sheaths | Opponens pollicis muscle | – | 194 | 384 |
| Muscles & sheaths | Palmar aponeurosis | – | 749 | 1284 |
| Muscles & | Palmar interossei muscles | – | 87 | 174 |

| | | | | |
|---|---|---|---|---|
| Muscles & sheaths | | | | |
| Muscles & sheaths | Palmaris longus muscle | – | 1618 | 3246 |
| Muscles & sheaths | Pronator quadratus | – | 527 | 1020 |
| Muscles & sheaths | Radial head of flexor digitorum superficialis | – | 510 | 1016 |
| Muscles & sheaths | Superficial head of flexor pollicis brevis | – | 136 | 276 |
| Muscles & sheaths | Superficial head of pronator teres | – | 258 | 516 |
| Muscles & sheaths | Superficial transverse metacarpal ligament | – | 162 | 192 |
| Muscles & sheaths | Supinator | – | 86 | 168 |
| Muscles & sheaths | Synovial sheaths of digits of hand | – | 3494 | 6976 |
| Muscles & sheaths | Tendon sheath - abd. pollicis longus - ext. pollicis brevis | – | 342 | 680 |
| Muscles & sheaths | Tendon sheath of extensor digitorum and extensor indicis | – | 1186 | 2368 |
| Muscles & sheaths | Tendon sheath of extensors carpi radialis | – | 1154 | 2304 |
| Muscles & sheaths | Transverse head of adductor pollicis | – | 509 | 1014 |
| Muscles & sheaths | Ulnar head of extensor carpi ulnaris | – | 255 | 506 |
| Muscles & sheaths | Ulnar head of flexor carpi ulnaris | – | 453 | 904 |
| Arteries | Common palmar digital arteries | – | 1632 | 3168 |
| Arteries | Deep palmar arch | FMA22839 | 1164 | 2304 |
| Arteries | Dorsal carpal anastomosis | – | 300 | 576 |
| Arteries | Dorsal carpal branch of ulnar artery | – | 876 | 1728 |
| Arteries | Dorsal digital arteries of hand | – | 7584 | 14976 |
| Arteries | Dorsal metacarpal arteries | – | 2940 | 5760 |
| Arteries | Palmar carpal branch of radial artery | – | 300 | 576 |
| Arteries | Palmar metacarpal arteries | – | 2208 | 4320 |
| Arteries | Posterior interosseous artery | – | 420 | 826 |
| Arteries | Proper palmar digital arteries | – | 15672 | 31104 |
| Arteries | Radial artery | FMA22733 | 1176 | 2338 |
| Arteries | Superficial palmar arch | FMA22835 | 1452 | 2880 |
| Arteries | Ulnar artery | FMA22797 | 504 | 994 |

| | | | | |
|---|---|---|---|---|
| Veins | Basilic vein | FMA22909 | 731 | 1448 |
| Veins | Cephalic vein | FMA13325 | 1079 | 2144 |
| Veins | Deep venous palmar arch | – | 2460 | 4896 |
| Veins | Dorsal digital veins of hand | – | 9324 | 18432 |
| Veins | Dorsal venous network of hand | – | 1308 | 2592 |
| Veins | Median antebrachial vein | FMA22968 | 1865 | 3678 |
| Veins | Palmar digital veins | – | 14412 | 28512 |
| Veins | Radial veins | – | 744 | 1474 |
| Veins | Superficial venous palmar arch | – | 2172 | 4320 |
| Veins | Ulnar veins | – | 936 | 1858 |
| Nerves | (Communicating branch of median nerve with ulnar nerve) | – | 300 | 576 |
| Nerves | Anterior branch of medial antebrachial cutaneous nerve | – | 180 | 346 |
| Nerves | Anterior interosseous nerve of forearm | – | 480 | 946 |
| Nerves | Common palmar digital branches of median nerve | – | 1188 | 2304 |
| Nerves | Common palmar digital branches of ulnar nerve | – | 900 | 1728 |
| Nerves | Deep branch of radial nerve | – | 492 | 970 |
| Nerves | Deep branch of ulnar nerve | – | 732 | 1440 |
| Nerves | Dorsal branch of ulnar nerve | – | 744 | 1440 |
| Nerves | Dorsal digital branches of radial nerve | – | 6852 | 13536 |
| Nerves | Dorsal digital branches of ulnar nerve | – | 2484 | 4896 |
| Nerves | Lateral antebrachial cutaneous nerve | – | 324 | 634 |
| Nerves | Median nerve | – | 576 | 1138 |
| Nerves | Muscular branches of ulnar nerve | – | 156 | 288 |
| Nerves | Palmar branch of median nerve | – | 588 | 1152 |
| Nerves | Palmar branch of ulnar nerve | – | 588 | 1152 |
| Nerves | Posterior antebrachial cutaneous nerve | – | 407 | 800 |
| Nerves | Posterior branch of medial antebrachial cutaneous nerve | – | 264 | 514 |
| Nerves | Posterior interosseous nerve of forearm | – | 420 | 826 |
| Nerves | Proper palmar digital branches of median nerve | – | 5556 | 10944 |
| Nerves | Proper palmar digital branches of ulnar nerve | – | 2916 | 5760 |
| Nerves | Superficial branch of radial nerve | – | 1776 | 3466 |
| Nerves | Superficial branch of ulnar nerve | – | 444 | 864 |
| Nerves | Ulnar nerve | – | 600 | 1186 |

**Supplementary Table S2 Modality comparison.**

| **Modality** | **Resolution (μm)** | **Depth (mm)** | **Capillary loops** | **Role** |
|---|---|---|---|---|

| | | | | |
|---|---|---|---|---|
| Conventional radiography (DR/CR) | 200 | – | No | Bone and joint |
| DSA | 200 | – | No | Arteries ≥200 µm |
| Phase-contrast / micro-CT | 1–100 | – | Ex vivo | Ex vivo / small animal |
| Nailfold VCP | 5 | 0.15 | Yes | Capillary morphology |
| OCT / OCTA | 7.5 | 1.5 | Yes | Retina; nailfold research |
| SDF / IDF | 1 | 0.05 | Yes | Sublingual microcirculation |
| Laser speckle contrast | 300 | 0.5 | Flow map | Perfusion |
| NIRS / diffuse optics | 10000 | 30 | No | Regional oxygenation |
| Photoacoustic microscopy | 5 | 1.0 | Yes | Preclinical / emerging |
| Ultrasound localization | 15 | 10 | Yes | Research / early clinical |

Resolution and penetration depth are order-of-magnitude values used in the physics module.

**Supplementary Table S3 Default model estimates.**

| Readout | Default | Unit |
|---|---|---|
| Capillary loop density | 8.0 | loops/mm |
| Erythrocyte velocity | 0.70 | mm/s |
| Capillary $SO_2$ | 86 | % |
| Tissue $PO_2$ | 36 | mmHg |
| Microvascular flow index | 3.0 | 0–3 |
| Heterogeneity index | 21 | % |
| Oxygen extraction ratio | 23 | % |
| Blood flow | 4.5 | mL/100 g/min |
| Capillary transit time | 0.85 | s |

All values are SIM model estimates.

**Supplementary Table S4 Simulated NVC profiles.**

| Pattern | Density | Giant | Avascular | Haem. | Tort. | Ramified | Severity |
|---|---|---|---|---|---|---|---|
| | (loops/mm) | (%) | (%) | (n) | (%) | (%) | |
| Healthy | 9.2 | 0 | 0 | 0.0 | 6 | 0 | 0.00 |
| Raynaud | 8.0 | 2 | 3 | 0.6 | 18 | 3 | 0.30 |
| SSc early | 7.4 | 8 | 6 | 1.5 | 16 | 6 | 0.45 |
| SSc active | 5.6 | 22 | 18 | 4.5 | 26 | 20 | 0.70 |
| SSc late | 3.0 | 6 | 45 | 1.0 | 34 | 42 | 0.95 |
| Diabetes | 6.7 | 3 | 14 | 1.2 | 55 | 10 | 0.60 |

Profile values at severity = 1.0; the interface and Fig. 5g show the blended values at the nominal severity. Fractions are proportions of the field or of loops; hemorrhages are expected counts.

**Supplementary Note**

**Source data and code.** The atlas geometry is derived from Z-Anatomy (CC BY-SA 4.0) and BodyParts3D (CC BY 4.0); the application code is MIT-licensed.

**Licenses and attribution.** Z-Anatomy: CC BY-SA 4.0 (repository) with BodyParts3D geometry (CC BY 4.0). Three.js r140: MIT. MediaPipe Tasks Vision: Apache-2.0. h264-mp4-encoder: MIT. The hand images are illustrative renderings prepared for this article and are not clinical images.

**Funding.** This work was supported by the Beijing Municipal Natural Science Foundation (Grant No. 7252093).
